# Free testosterone and malignant melanoma risk in men: prospective analyses of testosterone and SHBG with 19 cancers in men and postmenopausal women UK Biobank

**DOI:** 10.1101/2020.12.03.20241976

**Authors:** Eleanor L. Watts, Aurora Perez-Cornago, Anika Knuppel, Konstantinos K. Tsilidis, Timothy J. Key, Ruth C. Travis

## Abstract

We investigated the associations of estimated free and total circulating testosterone and sex hormone-binding globulin (SHBG) with cancer risk in men and postmenopausal women, using a pan-cancer approach, including 19 cancers in UK Biobank.

Risk was estimated using multivariable-adjusted Cox regression in up to 182,608 men and 122,112 postmenopausal women who were cancer-free at baseline. Participants diagnosed with cancer within two years of baseline were excluded. Hazard ratios (HRs) and confidence intervals (CIs) were corrected for regression dilution bias using repeat measurements. We accounted for multiple testing using the false discovery rate.

In men, higher free testosterone was associated with higher risks of melanoma and prostate cancer (HR per 50 pmol/L increase=1.35, 95% CI 1.14-1.61 and 1.10,1.04-1.18, respectively). Higher total testosterone was associated with an elevated risk of liver cancer (HR per 5 nmol/L=2.45,1.56-3.84), and higher SHBG was associated with a higher risk of liver cancer (HR per 10 nmol/L=1.56,1.31-1.87) and a lower risk of prostate cancer (0.93,0.91-0.96); associations with liver cancer were attenuated after excluding early follow-up. In postmenopausal women, higher free and total testosterone and lower SHBG were associated with elevated risks of endometrial (HR per 10 pmol/L=1.59,1.32-1.90; HR per 0.5 nmol/L=1.34,1.18-1.52 and HR per 25 nmol/L=0.78,0.67-0.91, respectively) and breast cancer (1.32,1.22-1.43;1.24,1.17-1.31 and 0.88,0.83-0.94, respectively).

We report a novel association of free testosterone with malignant melanoma in men; our findings also support known associations between sex hormones and risks for prostate, breast and endometrial cancers. The association with liver cancer in men may be attributable to reverse causation.

## Introduction

It is well-established that men have a higher risk of most non–sex-specific cancers than women(1,2). While some of these differences are due to lifestyle factors, there is also evidence that sex hormones are involved in the development of several cancers(3-6). However, aside from prostate(7-9), breast(9-11), and endometrial(9,12,13) cancers, previous population-based observational with hormone measurements have had limited power to assess associations with other cancer sites(4,5,14). Current evidence of associations with other cancers is largely based on animal models and tumor cell lines(15-17), or investigated indirectly via associations with self-reported menstrual and reproductive factors in women(18,19).

There are large sex differences in the regulation and quantities of serum androgens in men and postmenopausal women, therefore associations with disease risk should be assessed separately(20). In men, testosterone is primarily synthesised by the testes and serum concentrations of free testosterone are regulated by the hypothalamic-pituitary-gonadal (HPG) axis(21); while in women, testosterone is synthesised by the adrenal glands and ovaries, and the HPG axis is thought to have little direct effect in the regulation of testosterone concentrations(22). In both sexes, sex hormone-binding globulin (SHBG) is primarily synthesised by the liver and is secreted into the bloodstream, where it tightly binds to sex hormones and modulates their bioavailability(23).

In this paper, we aimed to examine the associations of serum concentrations of free testosterone, total testosterone and SHBG with the diagnosis of 19 cancers in a cohort of 182,600 men and 122,100 postmenopausal women in the UK Biobank, who were cancer-free >2 years from study baseline. This comprehensive pan-cancer approach, using standardised hormone and cancer endpoint analyses, enables the comparison of effect estimates and the exploration of the specificity of our findings.

## Materials and methods

### Study design

UK Biobank is a prospective cohort for public health research. Details of the study protocol and data collection are available online (http://www.ukbiobank.ac.uk/wp-content/uploads/2011/11/UK-Biobank-Protocol.pdf) and elsewhere(24,25).

In brief, all participants were registered with the UK National Health Service (NHS) and lived within 40 km of one of the UK Biobank assessment centres. Approximately 9.2 million people were initially invited to participate. Overall, 503,317 participants aged 40-69 years consented to join the cohort and attended one of 22 assessment centres throughout England, Wales and Scotland between 2006-2010, a participation rate of 5.5%(25).

The UK Biobank study was approved by the North West Multi-Centre Research Ethics Committee (reference number 06/MRE08/65), and at recruitment all participants gave written informed consent to participate and for their health to be followed-up through linkage to electronic medical records.

### Baseline assessment

At the baseline assessment visit, participants provided information on a range of sociodemographic, anthropometric, lifestyle, and health-related factors via a self-completed touch-screen questionnaire and a computer assisted personal interview(25).

### Blood sampling and biomarker assays

At recruitment, blood sampling was successfully performed in 99.7% of the cohort. Blood was collected in a serum separator tube and shipped to the central processing laboratory in temperature-controlled boxes at 4°C(26), then aliquoted and stored in a central working archive at −80°C(27). Serum concentrations of SHBG, testosterone and albumin were attempted in all participants. SHBG and testosterone (Beckman Coulter AU5800) were measured by chemiluminescent immunoassays, and albumin was measured by a colorimetric assay (Beckman Coulter AU5800). Average within-laboratory (total) coefficients of variation for low, medium, and high internal quality control level samples for each biomarker ranged from 2.1-8.3%.

Participants with total testosterone below the limit of detection were assigned a serum concentration of 0.26 nmol/L (3/4 of the lower limit of detection) (n=16 men and 25,253 postmenopausal women).

Serum insulin-like growth factor-I (IGF-I), glycated haemoglobin (HbA1c) and C-reactive protein (CRP) concentrations were also assayed in serum from blood collected at study baseline. Full details of the assay methods and quality assurance protocols are available online (https://biobank.ndph.ox.ac.uk/showcase/showcase/docs/serum_biochemistry.pdf).

### Free testosterone calculation

In the circulation, testosterone is bound to SHBG and albumin. Approximately 2% of total testosterone circulates unbound or “free” and is postulated to be biologically active(23). Free testosterone concentrations were estimated using a formula based on the law of mass action and measured total testosterone, SHBG and albumin concentrations(7,28).

### Repeat measurements

Repeat assessments were completed in 20,000 individuals. Participants who lived within a 35 km radius were invited via email to attend a repeat assessment clinic at the UK Biobank Co-ordinating Centre in Stockport between August 2012 and June 2013, with a response rate of 21%(29).

### Participant follow-up

Cancer registration data were provided via record linkage to the NHS Central Register and obtained via NHS Digital, until the censoring date (31^st^ March 2016 in England and Wales and 31^st^ October 2015 in Scotland).

Endpoints were defined as the first incident cancer diagnosis, or cancer recorded on death certificate if not previously diagnosed with cancer. Cancers were coded using the International Classification of Diseases Tenth revision code (ICD-10)(30) and we restricted analyses to cancers with a minimum of 100 recorded cases in men, and 100 cases for female only cancers (for consistency across sexes): lip, oral and pharynx (C00-14), oesophagus (C15), adenocarcinoma of oesophagus (C15, morphology codes ICD-O-3 8140-8573), stomach (C16), colorectum (C18-20) including colon (C18) and rectum (including rectosigmoid junction; C19-20), liver (C22), pancreas (C25), lung (C34), malignant melanoma (C43), mesothelioma (C45), female breast (C50), endometrial (C54.1), ovarian (C56), prostate (C61), kidney (C64-65), bladder (C67), brain (C71), non-Hodgkin lymphoma (NHL) (C82-85), multiple myeloma (C90), and leukemia (C91-95), and the NHL subtype diffuse NHL (C83). Person-years were calculated from the date of recruitment to the date of the first cancer registration, death, loss to follow-up, or censoring date, whichever occurred first.

### Exclusion criteria

Our analytical dataset included up to 182,608 men and 122,112 postmenopausal women (Supplementary Figure S1). We excluded 27,170 participants with prevalent cancer (except C44: non-melanoma skin cancer), 32,563 with no blood measurement data available or who had biomarker measurements that did not pass quality control procedures(31), 1,533 who were missing BMI measurement data and 22,697 who were diabetic at baseline (self-reported). We also excluded 6,070 participants who were diagnosed with cancer <2 years from baseline, 2,629 who were taking hormone-related medication at baseline, and 3,843 participants for whom it was not possible to determine genetic sex or whose genetic sex was identified as being different from their reported sex.

In women, we also excluded 66,526 participants who were either premenopausal or unknown (postmenopausal was defined as stopped period, aged >55 years), 12,745 women who were current users of hormone replacement therapy (HRT) or the contraceptive pill, 930 who had prevalent *in situ* breast cancer at study baseline, and 19,786 who reported having a hysterectomy or bilateral oophorectomy at study baseline (Supplementary Figure S1).

### Statistical analysis

Cox proportional hazards models were used to estimate the associations with cancer diagnosis, with age as the underlying time variable. Primary analyses were stratified by geographic area (10 UK regions) and age at recruitment (<45, 45-49, 50-54, 55-59, 60-64, ≥65 years), and adjusted for Townsend deprivation score (fifths, unknown (0.1%)), education level (college or university degree/vocational qualification, national examination at ages 17/18, national examination at age 16, other qualification or unknown (19.0%)), racial/ethnic group (white, Asian, black, mixed background and other, and unknown (1.2%)), height (<170, ≥170–<175, ≥175–<180, ≥180 cm, and unknown (0.1%), for men; and <160, ≥160– <165, ≥165–<170, ≥170 cm, and unknown (0.02%), for women), BMI (<25, ≥25–<30, ≥30– <35, ≥35 kg/m^2^), cigarette smoking (never, former, current light smoker (1-<15 cigarettes per day), current heavy smoker (≥15 cigarettes per day), current (number of cigarettes per day unknown), and smoking status unknown (0.5%)), alcohol consumption (non-drinkers, <1- <10, ≥10-<20, ≥20 g ethanol/day, unknown (0.6%)), and total physical activity (<10, 10-19, 20-39, 40-59, ≥60 metabolic equivalent of task hours per week, and unknown (0.6%)). For postmenopausal women, we additionally adjusted for past HRT use (never, ever), past oral contraceptive pill use (never, ever), parity and age at first birth (nulliparous; 1-2, <25; 1-2, 25-29; 1-2, ≥30; 1-2, unknown; ≥3, <25; ≥3, 25-29; ≥3, ≥30 years, unknown (16.0%)), age at menarche (<12, 12-13,≥14 years, unknown (2.9%)), and age at menopause (<45, 45-49, 50-54, ≥ 55 years, unknown (14.9%)). Adjustment covariates were defined *a priori* based on previous analyses of UK Biobank data(32).

Hazard ratios (HRs) and 95% confidence intervals (CIs) were estimated using the median value within each fourth and calibrated to allow risk estimates on a continuous scale (per 50 pmol/L, per 5 nmol/L and 10 nmol/L increments for free testosterone, total testosterone and SHBG, respectively for men and per 10 pmol/L, per 0.5 nmol/L and 25 nmol/L, respectively for postmenopausal women; these increments were chosen based on the sex-specific hormone standard deviations [SDs]).

Measurement error and within person variability using single measures at baseline leads to under-estimation of risk (i.e. regression dilution bias)(33). Estimates for trend were corrected for regression dilution bias by assigning the median values from the fourths of blood biomarker concentrations measured in the repeat blood sample for the subcohort who attended the repeat assessment (up to 7,669 men and 4,568 postmenopausal women, median of 4.3 years after first blood collection(34)) to women grouped by fourths of hormone concentrations measured at study baseline(35), excluding participants who were diagnosed with cancer between baseline and repeat assessment (up to 132 men and 48 women).

In the categorical analyses, biomarker measurements were categorised into fourths and HRs were calculated relative to the lowest fourth of each blood parameter. The variance of the log risk in each group was calculated (from the variances and covariances of the log risk) and used to obtain group-specific 95% CIs, which enable comparisons across different exposure categories(36).

### Sensitivity analyses

#### Subgroup analyses

Subgroup analyses for incident cancer were examined with participants categorised according to: i) time from blood collection to diagnosis (>2 -≤4.7, >4.7 years) ii) age at diagnosis (≤65, >65 years), and iii), age at blood collection (≤60, >60 years), categories were based on the median values within this sample. Heterogeneity in the associations for case-specific variables (i.e. time to diagnosis and age at diagnosis) was examined using stratified Cox models based on competing risks, comparing the risk coefficients and standard errors in the two subgroups, and testing with a χ^2^ for heterogeneity. For age at blood collection, heterogeneity was assessed using a χ^2^ interaction term.

#### Adjustment for other factors

To assess the role of other biomarkers, we further adjusted the primary analysis for serum IGF-I, HbA1c and CRP concentrations (fourths, unknown). The association of testosterone concentration and melanoma may have been confounded by factors relating to sun exposure and sensitivity, therefore we additionally adjusted for: skin color (very fair, fair, light olive, dark olive, brown/black, missing); hair color (blonde, red, light brown, dark brown, black, missing); skin reaction to sun exposure (get very tanned, moderately tanning, mildly/occasionally tanning, never tanning only burning, missing); and sunburn before age 15 (never, ever, missing).

All analyses were performed using Stata version 14.1 (Stata Corporation, College Station, TX, USA), and figures were plotted in R version 3.6.3. All tests of significance were two-sided, and P-values <0.05 were considered statistically significant. In the primary analysis, we additionally accounted for multiple testing using false discovery rates (FDR; 48 statistical tests in men and 54 statistical tests in women)(37).

## Results

After a mean follow-up of 7.0 years (SD=1), 9,519 men (5.2%) and 5,632 postmenopausal women (4.6%) were diagnosed with any type of malignant cancer (excluding non-melanoma skin cancer, C44).

Table 1 summarises the baseline characteristics of study participants. Mean age at recruitment was 56.1 (SD=8.2) in men and 60.2 years (5.3) in postmenopausal women. Participants who subsequently developed cancer were older, were more likely to smoke and had worse self-rated health at recruitment. Men who were subsequently diagnosed with cancer on average had a slightly lower socioeconomic status, while women who were diagnosed with cancer had a slightly higher socioeconomic status and were more likely to have previously used HRT compared those without a cancer diagnosis (Table 1). Mean and SD values for baseline biomarker measurements are displayed in Table 1.

**Table 1:** Baseline characteristics and blood data in men and postmenopausal women (in the analytic sample and incident cancers cases) in UK Biobank.

|  | Men |  | Postmenopausal women |  |
| --- | --- | --- | --- | --- |
|  | All (n=182,608) | Incident cancer*<br>(n=9,519) | All (n=122,112) | Incident cancer*<br>(n=5,632) |
| **Sociodemographic** |  |  |  |  |
| Age at recruitment (years), mean (SD) | 56.13 (8.22) | 61.07 (6.18) | 60.15 (5.29) | 61.29 (4.97) |
| Most deprived quintile, % (N) | 19.58 (35,716) | 19.02 (1,808) | 17.24 (21,037) | 19.04 (1,072) |
| Black ethnicity, % (N) | 1.38 (2,485) | 0.94 (88) | 1.02 (1,238) | 0.59 (33) |
| Not in paid/self-employment, % (N) | 36.52 (66,693) | 53.50 (5,093) | 54.93 (67,082) | 60.58 (3,412) |
| Living with partner, % (N) | 76.91 (140,452) | 77.08 (7,337) | 69.48 (84,849) | 67.86 (3,822) |
| **Anthropometric, mean (SD)** |  |  |  |  |
| Height (cm) | 175.76 (6.82) | 175.32 (6.70) | 161.92 (6.19) | 162.09 (6.32) |
| BMI (kg/m<sup>2</sup>) | 27.61 (4.05) | 27.69 (4.02) | 26.96 (4.90) | 27.53 (5.15) |
| **Lifestyle, % (N)** |  |  |  |  |
| Current cigarette smokers | 12.53 (22,744) | 15.14 (1,430) | 7.90 (9,610) | 10.77 (604) |
| Drinking alcohol $\geq 20$ g per day | 44.37 (80,618) | 46.38 (4,390) | 13.50 (16,378) | 14.61 (818) |
| Low physical activity (0-10 METs per week) | 27.32 (48,357) | 27.64 (2,545) | 30.83 (36,136) | 32.82 (1,770) |
| **Health history, % (N)** |  |  |  |  |
| Hypertension | 51.82 (94,587) | 59.27 (5,637) | 46.25 (56,422) | 50.11 (2,817) |
| Poor self-rated health | 3.95 (7,164) | 4.49 (425) | 2.76 (3,355) | 3.39 (190) |
| **Reproductive factors, % (N)** |  |  |  |  |
| Previous use of HRT | - | - | 42.56 (51,965) | 46.64 (2,627) |
| Previous use of oral contraceptive pill | - | - | 78.26 (95,570) | 75.30 (4,241) |
| Nulliparous | - | - | 15.99 (19,514) | 16.67 (938) |
| <25 years at first child | - | - | 55.06 (47,996) | 57.96 (2,301) |
| $\geq 14$ years at menarche | - | - | 36.80 (43,619) | 36.34 (1,984) |
| <50 years at menopause | - | - | 32.19 (33,439) | 29.64 (1,401) |
| **Baseline blood measures, mean (SD)** |  |  |  |  |
| Free testosterone (pmol/L) | 210.49 (59.54) | 199.55 (54.92) | 14.41 (10.13) | 15.25 (10.59) |
| Total testosterone (nmol/L) | 12.11 (3.63) | 11.97 (3.62) | 1.09 (0.63) | 1.14 (0.73) |
| SHBG (nmol/L) | 39.65 (16.51) | 42.34 (17.21) | 58.91 (26.51) | 57.24 (26.00) |
| Albumin (g/L) | 45.59 (2.58) | 45.06 (2.60) | 45.11 (2.53) | 44.85 (2.56) |

\* Diagnosed > 2 years from baseline
Abbreviations: BMI=body mass index; HRT= hormone replacement therapy; METs=metabolic equivalent of tasks; SD=standard deviation; SHBG=sex hormone binding globulin

### Associations between testosterone and SHBG concentrations and cancer diagnosis

#### Men

Serum free testosterone concentration was positively associated with risks of malignant melanoma and prostate cancer (HR per 50 pmol/L increment=1.35, 95% CI 1.14-1.61; *P*_*trend*_=0.0006, and 1.10, 1.04-1.18; *P*_*trend*_=0.002, respectively) and inversely associated with the risk of leukemia (0.77, 0.60-0.99; *P*_*trend*_=0.04) (Figure 1).

**Figure 1:**
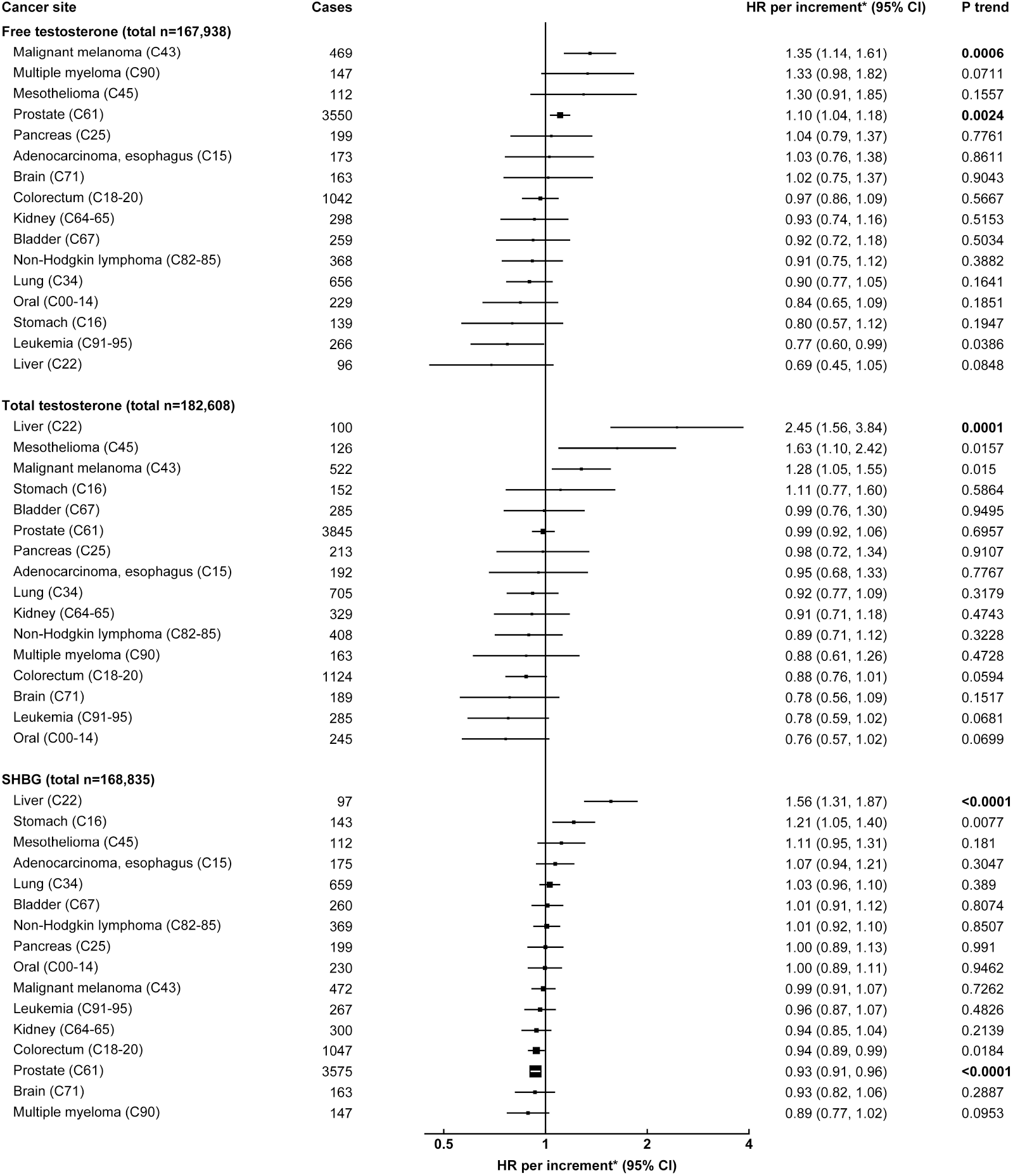
HRs and 95% CIs for cancer diagnosis per increment in free testosterone, total testosterone and SHBG concentrations by cancer site in men†. HRs are presented by squares with their 95% CIs as horizontal lines, the size of the squares is inversely proportional to the variance of the log HR. P_trends_ are bold if associations are statistically significant after accounting for false discovery rates. *Increments are free testosterone, per 50 pmol/L; total testosterone, per 5 nmol/L; SHBG, per 10 nmol/L. †Associations stratified for age group, geographical region and adjusted for Townsend deprivation score, racial/ethnic group, height, lives with a spouse or partner, body mass index, cigarette smoking, alcohol consumption, and total physical activity and corrected for regression dilution bias. Abbreviations: CI=Confidence interval; HR=hazard ratio; SHBG= sex hormone binding globulin.

Serum total testosterone concentration was positively associated with risks for liver cancer (HR per 5 nmol/L increment=2.45, 1.56-3.84; *P*_*trend*_=0.0001), mesothelioma (1.63, 1.10-2.42; *P*_*trend*_=0.02) and malignant melanoma (1.28, 1.05-1.55; *P*_*trend*_=0.02) (Figure 1).

Serum SHBG concentrations were positively associated with risks of liver (HR per 10 nmol/L increment=1.56, 1.31-1.87; *P*_*trend*_=<0.0001) and stomach cancer (1.21, 1.05-1.40; *P*_*trend*_=0.008) and inversely with risks of prostate (0.93, 0.91-0.96; *P*_*trend*_=<0.0001) and colorectal cancer (0.94, 0.89-0.99; *P*_*trend*_=0.02) (Figure 1).

After accounting for multiple testing, the associations of free testosterone with prostate cancer and melanoma, total testosterone with liver cancer, and SHBG with liver and prostate cancer remained statistically significant.

#### Postmenopausal women

Serum free testosterone concentration was positively associated with risks of endometrial and breast cancer (HR per 10 pmol/L increment=1.59, 1.32-1.90; *P*_*trend*_<0.0001, and 1.32, 1.22-1.43; *P*_*trend*_<0.0001, respectively), and inversely with risks of multiple myeloma (0.65, 0.43-0.97; *P*_*trend*_=0.03) and NHL (0.76, 0.59-0.99; *P*_*trend*_=0.04) (Figure 2).

**Figure 2:**
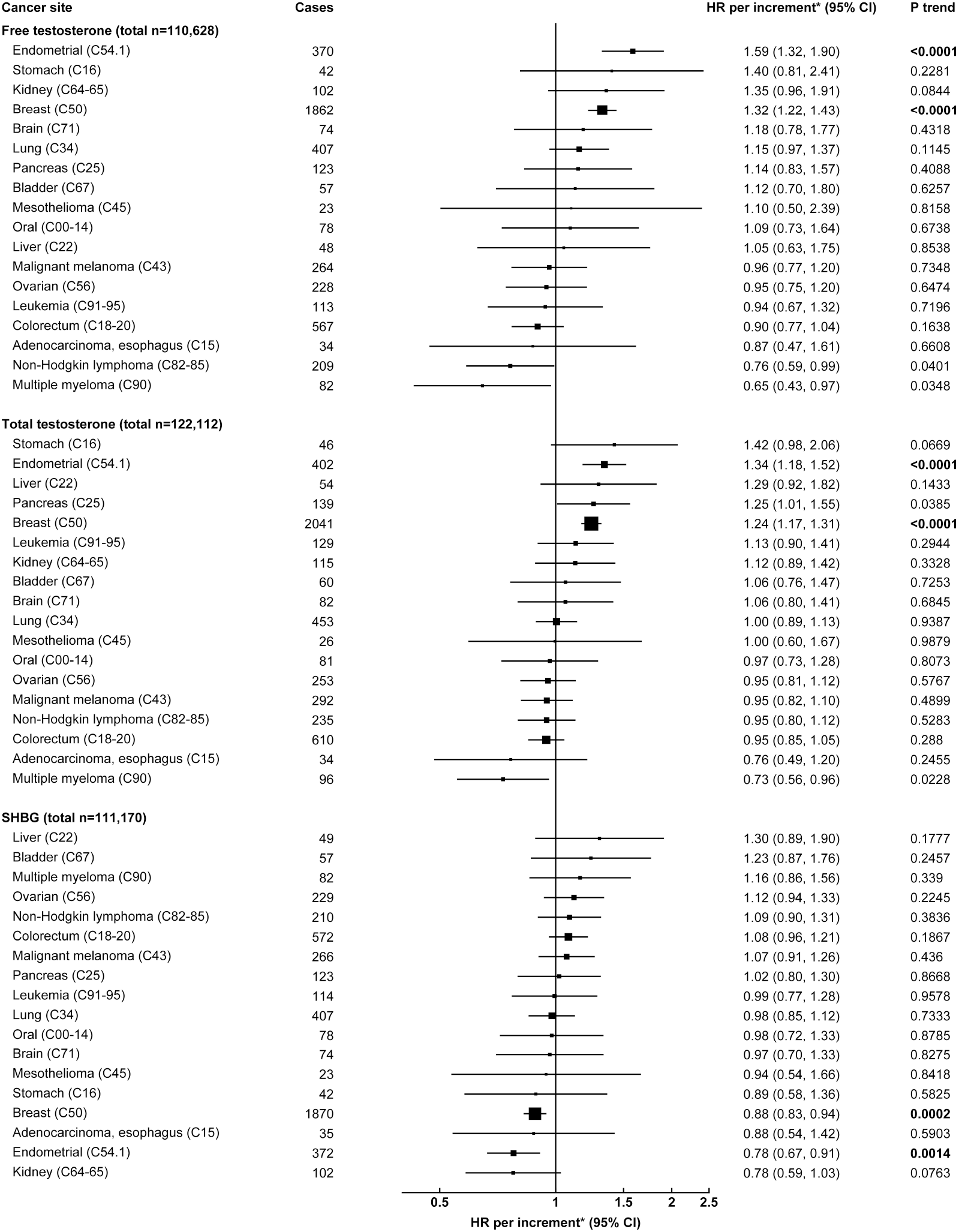
HRs and 95% CIs for cancer diagnosis per increment in free testosterone, total testosterone and SHBG concentrations by cancer site in postmenopausal women†. HRs are presented by squares with their 95% CIs as horizontal lines, the size of the squares is inversely proportional to the variance of the log HR. P_trends_ are bold if associations are statistically significant after accounting for false discovery rates. *Increments are free testosterone, per 10 pmol/L; total testosterone, per 0.5 nmol/L; SHBG, per 25 nmol/L. †Associations stratified for age group, geographical region and adjusted for Townsend deprivation score, racial/ethnic group, height, lives with a spouse or partner, body mass index, cigarette smoking, alcohol consumption, total physical activity, hormone replacement therapy use, oral contraceptive use, and parity and age at first birth, and corrected for regression dilution bias. Abbreviations: CI=Confidence interval; HR=hazard ratio; SHBG=sex hormone binding globulin.

Serum total testosterone concentration was positively associated with risks of endometrial (HR per 0.5 nmol/L increment=1.34, 1.18-1.52; *P*_*trend*_<0.0001), breast (1.24, 1.17-1.31; *P*_*trend*_<0.0001), and pancreatic cancer (1.25, 1.01-1.55; *P*_*trend*_=0.04) and inversely with risk of multiple myeloma (0.73. 0.56-0.96; *P*_*trend*_=0.02) (Figure 2).

Serum SHBG concentrations were inversely associated endometrial (HR per 25 nmol/L increment=0.78, 0.67-0.91; *P*_*trend*_=0.001) and breast cancer (0.88, 0.83-0.94; *P*_*trend*_=0.0002) (Figure 2).

After accounting for multiple testing, the associations of free and total testosterone and SHBG with endometrial and breast cancer remained statistically significant.

Results including cancer subtypes, fourths, adjustment models and associations with and without adjustment for regression dilution bias are displayed in Supplementary Tables S1-S6.

### Sensitivity analyses

#### Subgroup analyses

In men, the significant associations between testosterone and cancer diagnosis showed no significant heterogeneity by length of follow-up, age at diagnosis, or age at blood collection (Figure 3 and Supplementary Figures S2-3). There was evidence of heterogeneity in the association of SHBG with liver cancer by length of follow-up; men diagnosed with liver cancer 2-4.7 years from baseline had a higher risk of liver cancer diagnosis in relation to SHBG (HR per 10 nmol/L increment=1.92, 95% CI 1.45-2.55), while the magnitude of this association was smaller in men diagnosed >4.7 years from baseline (1.32, 1.05-1.67; P_het_=0.04).

**Figure 3:**
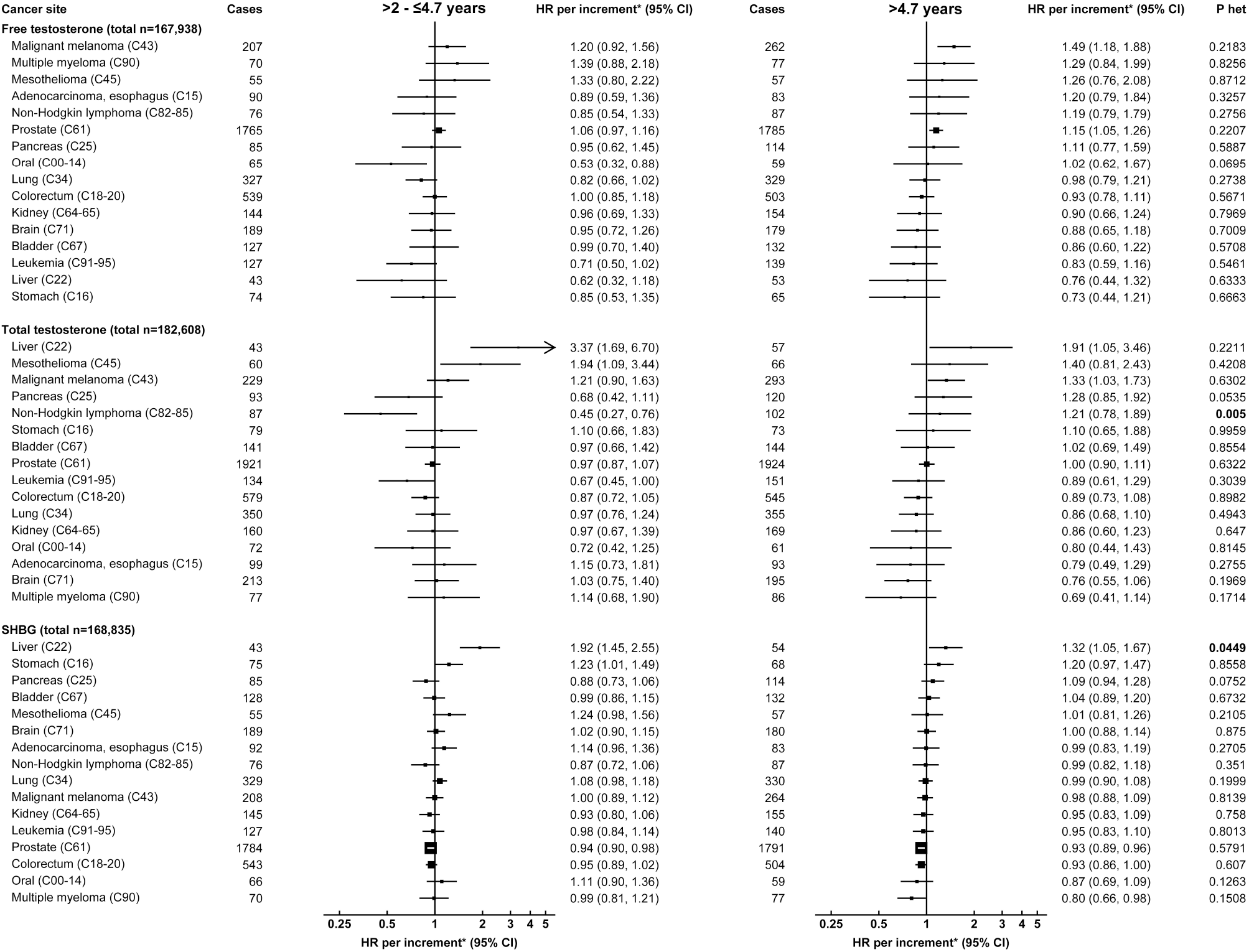
HRs and 95% CIs per increment in free testosterone, total testosterone and SHBG for cancer diagnosis by cancer site and time to diagnosis in men†. HRs are presented by squares with their 95% CIs as horizontal lines, the size of the squares is inversely proportional to the variance of the log HR. HRs are presented by squares with their 95% CIs as horizontal lines, the size of the squares is inversely proportional to the variance of the log HR. *P*_heterogeneity_ (Phet) comparing associations below and above time to diagnosis obtained using competing risks, p-values are bold if p<0.05. *Increments are free testosterone, per 50 pmol/L; total testosterone, per 5 nmol/L; SHBG, per 10 nmol/L. †Associations stratified for age group, geographical region and adjusted for Townsend deprivation score, racial/ethnic group, height, lives with a spouse or partner, body mass index, cigarette smoking, alcohol consumption and total physical activity. Follow-up time was based on the median time to diagnosis. Abbreviations: CI=Confidence interval; HR=hazard ratio; SHBG= sex hormone binding globulin.

In postmenopausal women, associations of testosterone and SHBG showed no significant heterogeneity by length of follow-up, age at diagnosis or age at blood collection (Figure 4 and Supplementary Figures S4-5), with the exception of total testosterone concentration and endometrial cancer by age at diagnosis and age at blood collection; postmenopausal women diagnosed with endometrial cancer aged ≤65 years had a larger magnitude of association of total testosterone with endometrial cancer risk (HR per 0.5 nmol/L increment=1.58, 95% CI 1.30-1.91), than women diagnosed aged >65 years (1.17, 0.98-1.39; P_het_=0.02) (Supplementary Figure 4). Similarly, women aged ≤60 years at blood collection had a larger magnitude of association (1.67, 1.32-2.12), than women >60 years at baseline (1.22, 1.05-1.42; P_het_=0.03).

**Figure 4:**
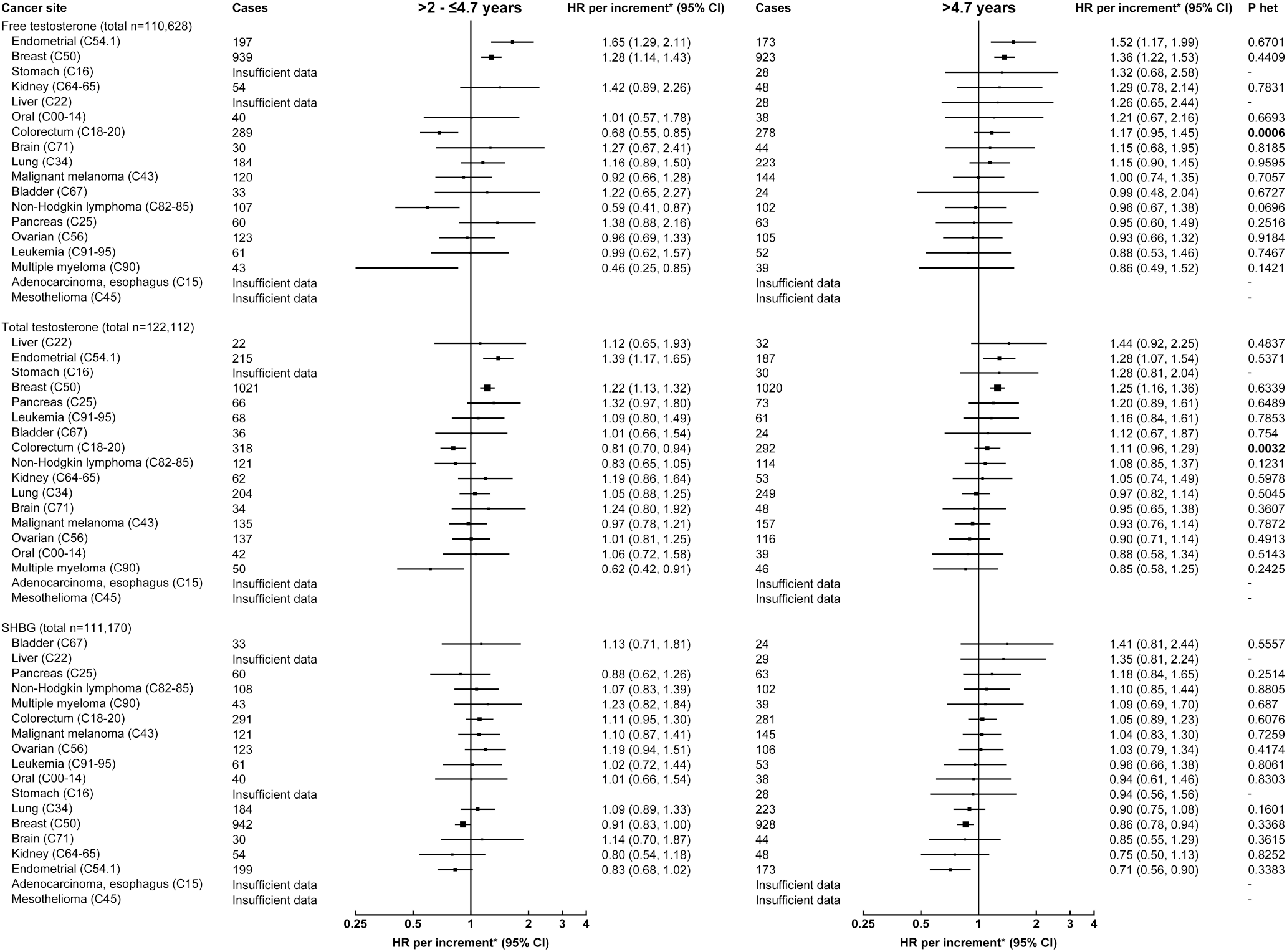
HRs and 95% CIs per free testosterone, total testosterone and SHBG increment for cancer diagnosis by cancer site and time to diagnosis in postmenopausal women†. HRs are presented by squares with their 95% CIs as horizontal lines, the size of the squares is inversely proportional to the variance of the log HR. HRs are presented by squares with their 95% CIs as horizontal lines, the size of the squares is inversely proportional to the variance of the log HR. *P*_heterogeneity_ (Phet) comparing associations below and above time to diagnosis obtained using competing risks, p-values are bold if p<0.05. *Increments are free testosterone, per 10 pmol/L; total testosterone, per 0.5 nmol/L; SHBG, per 25 nmol/L. †Associations stratified for age group, geographical region and adjusted for Townsend deprivation score, racial/ethnic group, height, lives with a spouse or partner, body mass index, cigarette smoking, alcohol consumption, total physical activity, hormone replacement therapy use, oral contraceptive use, and parity and age at first birth. Follow-up time was based on the median time to diagnosis. Abbreviations: CI=Confidence interval; HR=hazard ratio; SHBG= sex hormone binding globulin.

#### Adjustment for other factors

In men, following further adjustment for IGF-I, HbA1c and CRP, the association of SHBG concentration with colorectal cancer slightly attenuated (HR per 10 nmol/L increment=0.95, 95% CI 0.90-1.00; *P*_*trend*_=0.07) (Supplementary Table S4).

In postmenopausal women, further adjustment for these biomarkers slightly attenuated the association of serum free testosterone with NHL (HR per 10 pmol/L increment=0.78, 95% CI=0.60-1.01; *P*_*trend*_=0.06) (Supplementary Table S5).

The associations of free and total testosterone with melanoma in men were not materially different following further adjustment for sun exposure and sensitivity factors (Supplementary Table S7).

## Discussion

This comprehensive analysis of serum free and total testosterone and SHBG concentrations with cancer risk in 182,600 men and 122,100 postmenopausal women shows a novel association of free testosterone with melanoma risk in men. We also confirm previously observed associations with prostate(7-9), breast(9-11), and endometrial(9,12,13) cancer. We additionally report several possible associations with other cancer sites, particularly with SHBG and stomach cancer, but these associations were not statistically significant after accounting for multiple testing.

### Associations with incident cancer in men

Higher total and free testosterone concentrations were associated with an increased risk of malignant melanoma. Until now, associations between serum sex hormone concentrations and melanoma have not been robustly assessed in the context of other large prospective cohort studies (n=41 male melanoma cases)(14). However, it has been observed that men diagnosed with melanoma have a higher risk of developing prostate cancer and *vice versa*, suggesting a common biological cause(38). There is also biological evidence of plausibility; androgens are involved in several skin processes including melanogenesis(15,39); and *in vitro* models implicate a role of androgen signalling in promoting melanoma(15).

We have previously observed associations between free testosterone and prostate cancer risk and an inverse association with SHBG in UK Biobank, and in an international consortium of 20 cohorts (6,933 cases and 12,088 controls)(7,8). These associations are consistent with evidence from two large randomised controlled trials (which aimed to reduce intra-prostatic androgen signalling)(40,41) and a Mendelian randomization study(9). However, testosterone is a determinant of prostate-specific antigen concentrations(42), which are commonly used in the detection of prostate cancer, therefore whether the risk associations might be partly explained by detection bias remains under debate(43,44).

We note the large magnitude of the associations between total testosterone and SHBG and liver cancer, which has been observed previously(3). The liver is integral in sex hormone signalling and metabolism, and synthesis of SHBG. Liver damage often leads to abnormal concentrations of blood biomarkers. For instance, liver fibrosis, which can develop into liver cancer, is associated with higher SHBG concentrations in men(45). Therefore, higher SHBG concentration may be a marker of preclinical disease, which is supported by a marked attenuation of risk estimates when restricting cases to those diagnosed after the median follow-up.

We also observed nominally significant inverse associations between free testosterone and leukemia, total testosterone and mesothelioma, and SHBG and colorectal cancer, and a positive association between SHBG and stomach cancer risk. However, these associations did not withstand correction for multiple testing. While current evidence of the inverse associations between testosterone and/or SHBG and these cancers is limited, associations between sex hormones and stomach cancer risk have been observed previously(4). Speculatively, this may be due to the role of SHBG in limiting estradiol bioactivity, as estradiol has been shown to reduce the viability and carcinogenic potential of *H. pylori* in animal models(46,47). Nevertheless, we are unable to rule-out the possibility of reverse causation(48) or chance.

### Associations with incident cancer in postmenopausal women

In postmenopausal women, we observed positive associations of free and total testosterone with risks for endometrial and breast cancer, while higher SHBG was associated with a lower risk of cancer at these sites. These associations have been observed previously in prospective nested case-control studies(10,12,13) and in Mendelian randomization analyses(9). Associations with breast cancer have also been reported in pooled analyses by an international consortium of 15 cohorts with 3,112 cases and 6,117 controls(11). Higher androgen concentrations might increase the risk of these cancers indirectly, via their conversion to estrogens, which stimulate cell proliferation, increasing the probability of mutations and uncontrolled cell growth(49-51). There is also some evidence that testosterone may affect endometrial and breast cancer risk via androgen receptor signalling(52,53). In epidemiological analyses, when estimates of testosterone and breast cancer risk additionally adjusted for circulating estradiol concentrations the associations somewhat attenuated but remained statistically significant(10), while associations with endometrial cancer attenuated to the null(12,13).

We also observed possible inverse associations of testosterone with multiple myeloma, NHL and pancreatic cancer in postmenopausal women. However, these associations did not withstand correction for multiple testing.

### Differences in cancer risk in men vs postmenopausal women

Although men have a greater risk of most cancers in comparison with women(1,2), in this analysis, differences in testosterone and SHBG did not seem to account for this. Men were more likely to smoke, drink alcohol and have hypertension than postmenopausal women, which may indicate a greater importance of sex-differences in lifestyle factors in risk disparities, or differences in risk may be related to other sexual dimorphisms which are independent of sex hormones(2), including greater body size(54).

We did, however, observe a sex-specific association of free testosterone with melanoma. It is possible that these differences are due to the more than ten-fold greater free testosterone concentrations in men. National surveillance data indicate that women have a greater risk of melanoma from the age of puberty until ∼40 years old, and from the age of ∼50 men have a higher risk(55). This may indicate a relatively greater significance of hormones in premenopausal women, when sex hormone concentrations are markedly different, particularly estradiol, or the importance of other factors such as UV exposure. Large studies or pooling projects which include younger women are needed to investigate this further.

### Strengths and limitations

UK Biobank is currently the largest prospective study with hormone data available for nearly the whole cohort, providing a unique opportunity to robustly assess associations of serum testosterone and SHBG concentrations with cancer risk using a comprehensive pan-cancer approach. This greater power improved the precision of our risk estimates and enabled us to investigate associations with less common cancers, and the availability of repeat measurements enabled us to adjust for regression dilution bias(33). UK Biobank is also a well-characterised study population, and therefore we were able to adjust our risk estimates for a wide range of possible confounders and examine the role of other related biomarkers.

Limitations of the analysis are that tumor characterisation information is not currently available, which may be relevant for the etiology of some cancer sites. We were also underpowered to investigate heterogeneity in the associations, particularly for the less common cancers. The UK Biobank participants are predominantly white and healthier than the sampling population, therefore risk estimates may not be generalizable(25), though this is unlikely to affect the direction of the associations(56). Although all our analyses excluded participants diagnosed with cancer in the first 2 years of follow-up, the follow-up period is not very long and we are not able to completely rule-out the possibility of reverse causation. Although we employed FDR to address the increased risk of chance findings due to the large number of statistical tests, we cannot rule out the possibility of chance findings, particularly in the sensitivity analyses.

SHBG also modulates the bioavailability of other sex hormones including estradiol, which is correlated with testosterone and may independently affect cancer risk via estrogen receptor signalling. However, estradiol concentrations are too low in men and postmenopausal women to obtain accurate data with the assays used in the UK Biobank. Furthermore, 13.8% of postmenopausal women had testosterone concentrations below the limit of detection, this might reduce power and attenuate risk estimates. Free testosterone concentrations were estimated using a commonly-employed formula derived from mass action equations, however the extent to which free testosterone represents the biologically active component of total testosterone remains under debate(57).

In conclusion, our study reports a novel positive association of free testosterone with melanoma in men. Our findings also support established associations of serum testosterone concentrations with prostate cancer risk in men, and with breast and endometrial cancer risk in postmenopausal women. Associations with testosterone were sex and site-specific and therefore do not support a general role of testosterone in cancer risk. Studies with longer follow-up time and pooled datasets including genetic information are needed to further assess associations.

## Data Availability

All bona fide researchers can apply to use the UK Biobank resource for health-related research that is in the public interest (https://www.ukbiobank.ac.uk/register-apply/).

## Acknowledgements

We would like to thank Georgina K. Fensom for her contributions to the data analysis. We thank all participants, researchers, and support staff who made the study possible.

## Data availability

This research has been conducted using the UK Biobank Resource under application numbers 3282 and 24494. All bona fide researchers can apply to use the UK Biobank resource for health-related research that is in the public interest (https://www.ukbiobank.ac.uk/register-apply/).

