## Supplementary materials for "Free testosterone and malignant melanoma risk in men: prospective analyses of testosterone and SHBG with 19 cancers in men and postmenopausal women UK Biobank"

**Table of contents**

**Figure S1:** Participant exclusion criteria...………………………...………….…...……………...…….Page 2

Figure S2: HRs and 95% CIs per free testosterone, total testosterone and SHBG increment for cancer diagnosis by cancer site and age at diagnosis in men†…………………………………………………..Page 3

**Figure S3:** HRs and 95% CIs per free testosterone, total testosterone and SHBG increment for cancer diagnosis by cancer site and age at blood collection in men†………………………...…………………Page 4

**Figure S4:** HRs and 95% CIs per free testosterone, total testosterone and SHBG increment for cancer diagnosis by cancer site and age at diagnosis in postmenopausal women†…….……………………….Page 5

**Figure S5:** HRs and 95% CIs per free testosterone, total testosterone and SHBG increment for cancer diagnosis by cancer site and age at blood collection in postmenopausal women†……….……………..Page 6

Table S1: HR and 95% CIs for cancer diagnosis by serum free testosterone concentrations in men…..Page 7

Table S2: HR and 95% CIs for cancer diagnosis by serum total testosterone concentrations in men……………………………………………………………………………………………………..Page 10

Table S3: HR and 95% CIs for cancer diagnosis by serum SHBG concentrations in men………...…Page 13

**Table S4:** HR and 95% CIs for cancer diagnosis by serum free testosterone concentrations in postmenopausal women………………………………………………………………………………...Page 16

**Table S5:** HR and 95% CIs for cancer diagnosis by serum total testosterone concentrations in postmenopausal women……………………………………………………………………………...…Page 19

**Table S6:** HR and 95% CIs for cancer diagnosis by serum SHBG concentrations in postmenopausal women………………………………………………………………………………………………….Page 22

**Table S7:** HR and 95% CIs for malignant melanoma in men after correction for factors relating to sun exposure………………………………………………………………………………………………...Page 25

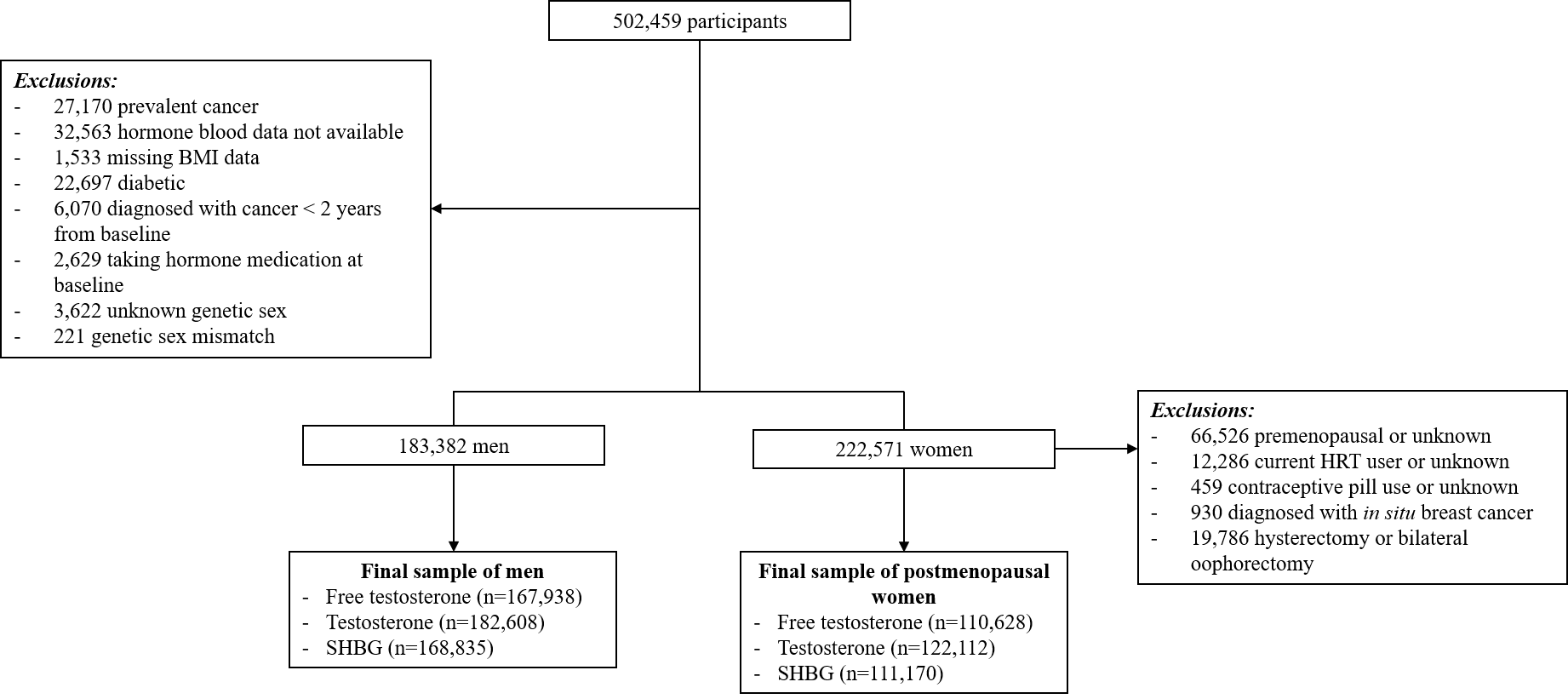

Supplementary Figure S1: Participant selection criteria

Abbreviations: BMI=body mass index, HRT=hormone replacement therapy, SHBG=sex hormone binding globulin.

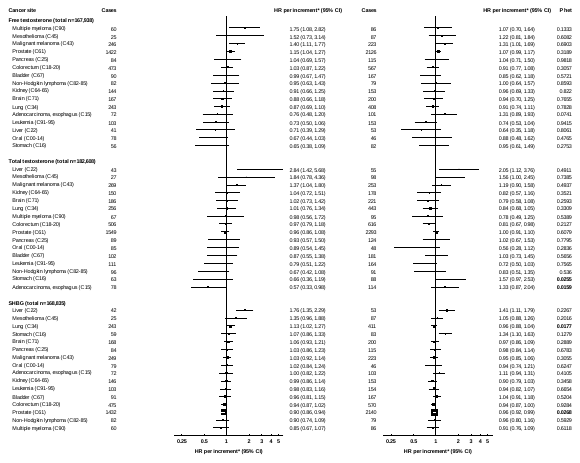

**> 65 years**

**≤ 65 years**

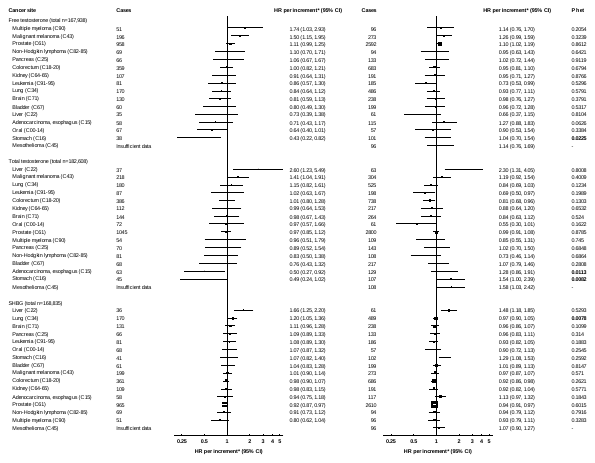

Supplementary Figure S2: HRs and 95% CIs per free testosterone, total testosterone and SHBG increment for cancer diagnosis by cancer site and age at diagnosis in men†

HRs are presented by squares with their 95% CIs as horizontal lines, the size of the squares is inversely proportional to the variance of the log HR. HRs are presented by squares with their 95% CIs as horizontal lines, the size of the squares is inversely proportional to the variance of the log HR. *P*_heterogeneity_ (Phet) comparing associations below and above age at diagnosis obtained using competing risks, p-values are bold if p<0.05.

*Increments are free testosterone, per 50 pmol/L; total testosterone, per 5 nmol/L; SHBG, per 10 nmol/L.

†Associations stratified for age group, geographical region and adjusted for Townsend deprivation score, racial/ethnic group, height, lives with a spouse or partner, body mass index, cigarette smoking, alcohol consumption and total physical activity. Age at diagnosis cut-point was based on the median value.

Abbreviations: CI=Confidence interval; HR=hazard ratio; SHBG= sex hormone binding globulin.

**> 60 years**

**≤ 60 years**

Supplementary Figure S3: HRs and 95% CIs per free testosterone, total testosterone and SHBG increment for cancer diagnosis by cancer site and age at blood collection in men†

HRs are presented by squares with their 95% CIs as horizontal lines, the size of the squares is inversely proportional to the variance of the log HR. HRs are presented by squares with their 95% CIs as horizontal lines, the size of the squares is inversely proportional to the variance of the log HR. *P*_heterogeneity_ (Phet) was assessed using a χ^2^ interaction term, p-values are bold if p<0.05.

*Increments are free testosterone, per 50 pmol/L; total testosterone, per 5 nmol/L; SHBG, per 10 nmol/L.

†Associations stratified for age group, geographical region and adjusted for Townsend deprivation score, racial/ethnic group, height, lives with a spouse or partner, body mass index, cigarette smoking, alcohol consumption and total physical activity. Age at blood collection cut-point was based on the median value.

Abbreviations: CI=Confidence interval; HR=hazard ratio; SHBG= sex hormone binding globulin.

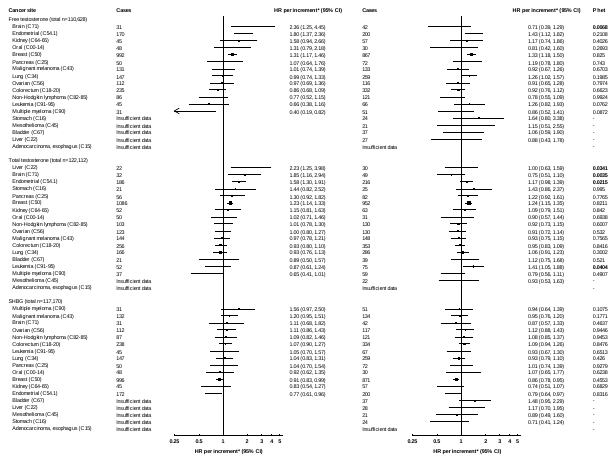

**> 65 years**

**≤ 65 years**

Supplementary Figure S4: HRs and 95% CIs per free testosterone, total testosterone and SHBG increment for cancer diagnosis by cancer site and age at diagnosis in postmenopausal women†

HRs are presented by squares with their 95% CIs as horizontal lines, the size of the squares is inversely proportional to the variance of the log HR. HRs are presented by squares with their 95% CIs as horizontal lines, the size of the squares is inversely proportional to the variance of the log HR. *P*_heterogeneity_ (Phet) comparing associations below and above age at diagnosis obtained using competing risks, p-values are bold if p<0.05.

*Increments are free testosterone, per 10 pmol/L; total testosterone, per 0.5 nmol/L; SHBG, per 25 nmol/L.

†Associations stratified for age group, geographical region and adjusted for Townsend deprivation score, racial/ethnic group, height, lives with a spouse or partner, body mass index, cigarette smoking, alcohol consumption, total physical activity, hormone replacement therapy use, oral contraceptive use, and parity and age at first birth. Age at diagnosis cut-point was based on the median value.

Abbreviations: CI=Confidence interval; HR=hazard ratio; SHBG= sex hormone binding globulin.

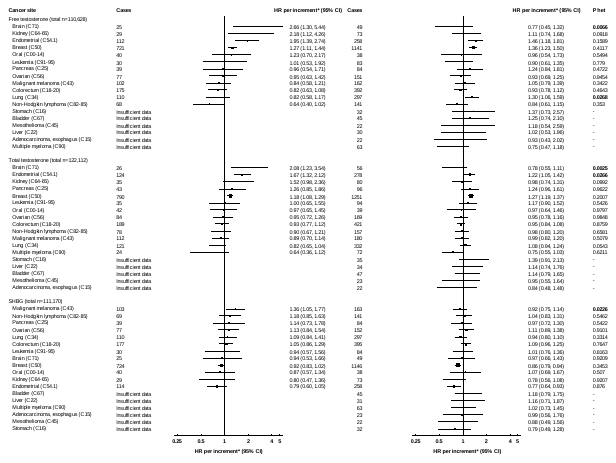

**> 60 years**

**≤ 60 years**

Supplementary Figure S5: HRs and 95% CIs per free testosterone, total testosterone and SHBG increment for cancer diagnosis by cancer site and age at blood collection in postmenopausal women†

HRs are presented by squares with their 95% CIs as horizontal lines, the size of the squares is inversely proportional to the variance of the log HR. HRs are presented by squares with their 95% CIs as horizontal lines, the size of the squares is inversely proportional to the variance of the log HR. *P*_heterogeneity_ (Phet) was assessed using a χ^2^ interaction term, p-values are bold if p<0.05.

*Increments are free testosterone, per 10 pmol/L; total testosterone, per 0.5 nmol/L; SHBG, per 25 nmol/L.

†Associations stratified for age group, geographical region and adjusted for Townsend deprivation score, racial/ethnic group, height, lives with a spouse or partner, body mass index, cigarette smoking, alcohol consumption, total physical activity, hormone replacement therapy use, oral contraceptive use, and parity and age at first birth. Age at blood collection cut-point was based on the median value.

Abbreviations: CI=Confidence interval; HR=hazard ratio; SHBG= sex hormone binding globulin.

Supplementary Table S1: HR and 95% CIs for cancer diagnosis by serum free testosterone concentrations in men

|  | Fourths of free testosterone | | | |  | per 50 pmol/L increment | | | |
| --- | --- | --- | --- | --- | --- | --- | --- | --- | --- |
|  |  |  |  |  |  | Not corrected for regression dilution bias | | Corrected for regression dilution bias | |
|  | 1 | 2 | 3 | 4 |  |  | P_trend_ |  | P_trend_ |
| Whole cohort, n | 41985 | 41984 | 41985 | 41984 |  | 167938 | | | |
| Median (IQR), pmol/L | 150.1 (28.4) | 188.1 (16.2) | 221.7 (19.0) | 274.1 (46.6) |  |  |  |  |  |
| Oral (C00-14) |  |  |  |  |  |  |  |  |  |
| Cases | 67 | 65 | 51 | 46 |  | 229 | | | |
| Model 0, HR (95% CI) | 1.00 (0.78-1.28) | 1.00 (0.78-1.27) | 0.81 (0.61-1.06) | 0.77 (0.57-1.04) |  | 0.89 (0.76-1.03) | 0.1175 | 0.82 (0.64-1.06) | 0.1280 |
| Model 1, HR (95% CI) | 1.00 (0.78-1.28) | 1.03 (0.81-1.31) | 0.85 (0.64-1.12) | 0.79 (0.59-1.07) |  | 0.90 (0.77-1.05) | 0.1730 | 0.84 (0.65-1.09) | 0.1851 |
| Model 2, HR (95%CI) | 1.00 (0.78-1.28) | 1.05 (0.83-1.34) | 0.87 (0.66-1.15) | 0.82 (0.61-1.11) |  | 0.91 (0.78-1.06) | 0.2337 | 0.86 (0.67-1.11) | 0.2482 |
| Esophagus (C15) |  |  |  |  |  |  |  |  |  |
| Cases | 67 | 55 | 55 | 39 |  | 216 | | | |
| Model 0, HR (95% CI) | 1.00 (0.78-1.28) | 0.96 (0.74-1.25) | 1.12 (0.86-1.45) | 1.04 (0.76-1.44) |  | 1.03 (0.88-1.21) | 0.6802 | 1.05 (0.81-1.36) | 0.7115 |
| Model 1, HR (95% CI) | 1.00 (0.78-1.28) | 1.03 (0.79-1.34) | 1.24 (0.95-1.61) | 1.18 (0.86-1.63) |  | 1.09 (0.93-1.27) | 0.2888 | 1.14 (0.88-1.49) | 0.3103 |
| Model 2, HR (95%CI) | 1.00 (0.78-1.28) | 1.04 (0.80-1.35) | 1.25 (0.96-1.63) | 1.19 (0.86-1.65) |  | 1.09 (0.93-1.28) | 0.2682 | 1.15 (0.89-1.50) | 0.2893 |
| Adenocarcinoma, esophagus (C15) | |  |  |  |  |  |  |  |  |
| Cases | 59 | 45 | 41 | 28 |  | 173 | | | |
| Model 0, HR (95% CI) | 1.00 (0.77-1.30) | 0.89 (0.67-1.19) | 0.95 (0.70-1.29) | 0.85 (0.58-1.24) |  | 0.94 (0.79-1.13) | 0.5324 | 0.91 (0.67-1.22) | 0.5168 |
| Model 1, HR (95% CI) | 1.00 (0.77-1.30) | 0.97 (0.73-1.30) | 1.09 (0.80-1.48) | 1.01 (0.69-1.49) |  | 1.02 (0.85-1.22) | 0.8390 | 1.03 (0.76-1.38) | 0.8611 |
| Model 2, HR (95%CI) | 1.00 (0.77-1.31) | 0.98 (0.73-1.31) | 1.09 (0.80-1.48) | 1.01 (0.69-1.49) |  | 1.02 (0.85-1.22) | 0.8328 | 1.03 (0.76-1.39) | 0.8553 |
| Stomach (C15) |  |  |  |  |  |  |  |  |  |
| Cases | 53 | 35 | 32 | 19 |  | 139 | | | |
| Model 0, HR (95% CI) | 1.00 (0.76-1.32) | 0.76 (0.55-1.06) | 0.81 (0.58-1.15) | 0.62 (0.39-0.99) |  | 0.84 (0.69-1.03) | 0.0949 | 0.74 (0.53-1.05) | 0.0882 |
| Model 1, HR (95% CI) | 1.00 (0.75-1.32) | 0.81 (0.58-1.12) | 0.88 (0.62-1.24) | 0.69 (0.43-1.09) |  | 0.88 (0.71-1.08) | 0.2071 | 0.80 (0.57-1.12) | 0.1947 |
| Model 2, HR (95%CI) | 1.00 (0.75-1.33) | 0.82 (0.59-1.14) | 0.90 (0.63-1.27) | 0.71 (0.44-1.12) |  | 0.89 (0.72-1.09) | 0.2516 | 0.81 (0.58-1.15) | 0.2366 |
| Colorectum (C18-20) |  |  |  |  |  |  |  |  |  |
| Cases | 344 | 266 | 243 | 189 |  | 1042 | | | |
| Model 0, HR (95% CI) | 1.00 (0.90-1.11) | 0.88 (0.78-1.00) | 0.93 (0.82-1.06) | 0.93 (0.80-1.07) |  | 0.97 (0.91-1.05) | 0.4613 | 0.95 (0.85-1.08) | 0.4435 |
| Model 1, HR (95% CI) | 1.00 (0.90-1.12) | 0.89 (0.79-1.00) | 0.95 (0.83-1.07) | 0.94 (0.81-1.09) |  | 0.98 (0.91-1.05) | 0.5893 | 0.97 (0.86-1.09) | 0.5667 |
| Model 2, HR (95%CI) | 1.00 (0.90-1.12) | 0.89 (0.79-1.00) | 0.94 (0.83-1.07) | 0.93 (0.80-1.08) |  | 0.98 (0.91-1.05) | 0.5215 | 0.96 (0.85-1.08) | 0.5011 |
| Colon (C18) |  |  |  |  |  |  |  |  |  |
| Cases | 222 | 156 | 149 | 114 |  | 641 | | | |
| Model 0, HR (95% CI) | 1.00 (0.87-1.14) | 0.80 (0.69-0.94) | 0.88 (0.75-1.03) | 0.86 (0.71-1.04) |  | 0.95 (0.86-1.04) | 0.2424 | 0.91 (0.78-1.06) | 0.2260 |
| Model 1, HR (95% CI) | 1.00 (0.87-1.15) | 0.82 (0.70-0.96) | 0.91 (0.77-1.07) | 0.90 (0.74-1.08) |  | 0.96 (0.88-1.06) | 0.4273 | 0.94 (0.80-1.09) | 0.4011 |
| Model 2, HR (95%CI) | 1.00 (0.87-1.15) | 0.82 (0.70-0.96) | 0.90 (0.77-1.06) | 0.89 (0.74-1.07) |  | 0.96 (0.87-1.05) | 0.3894 | 0.93 (0.80-1.09) | 0.3652 |
| Rectum (C19-20) |  |  |  |  |  |  |  |  |  |
| Cases | 123 | 112 | 96 | 75 |  | 406 | | | |
| Model 0, HR (95% CI) | 1.00 (0.83-1.20) | 1.04 (0.87-1.26) | 1.04 (0.85-1.27) | 1.03 (0.82-1.30) |  | 1.01 (0.90-1.13) | 0.8433 | 1.02 (0.84-1.23) | 0.8428 |
| Model 1, HR (95% CI) | 1.00 (0.83-1.20) | 1.03 (0.86-1.24) | 1.03 (0.84-1.25) | 1.01 (0.80-1.28) |  | 1.00 (0.89-1.13) | 0.9451 | 1.01 (0.83-1.22) | 0.9451 |
| Model 2, HR (95%CI) | 1.00 (0.83-1.20) | 1.03 (0.86-1.24) | 1.02 (0.84-1.25) | 1.00 (0.79-1.27) |  | 1.00 (0.89-1.12) | 0.9964 | 1.00 (0.83-1.21) | 0.9958 |
| Liver (C22) |  |  |  |  |  |  |  |  |  |
| Cases | 35 | 29 | 22 | 10 |  | 96 | | | |
| Model 0, HR (95% CI) | 1.00 (0.71-1.40) | 0.94 (0.65-1.35) | 0.81 (0.54-1.24) | 0.46 (0.24-0.86) |  | **0.76 (0.60-0.98)** | **0.0351** | **0.64 (0.42-0.97)** | **0.0341** |
| Model 1, HR (95% CI) | 1.00 (0.71-1.41) | 1.00 (0.70-1.44) | 0.90 (0.59-1.37) | 0.51 (0.27-0.96) |  | 0.80 (0.63-1.03) | 0.0876 | 0.69 (0.45-1.05) | 0.0848 |
| Model 2, HR (95%CI) | 1.00 (0.71-1.42) | 1.08 (0.75-1.54) | 1.01 (0.66-1.54) | 0.60 (0.32-1.12) |  | 0.86 (0.67-1.10) | 0.2286 | 0.77 (0.51-1.17) | 0.2216 |
| Pancreas (C25) |  |  |  |  |  |  |  |  |  |
| Cases | 69 | 42 | 51 | 37 |  | 199 | | | |
| Model 0, HR (95% CI) | 1.00 (0.79-1.27) | 0.70 (0.51-0.94) | 0.99 (0.76-1.31) | 0.96 (0.69-1.33) |  | 1.01 (0.86-1.19) | 0.9114 | 1.00 (0.77-1.32) | 0.9741 |
| Model 1, HR (95% CI) | 1.00 (0.78-1.28) | 0.71 (0.53-0.96) | 1.03 (0.78-1.36) | 1.01 (0.72-1.40) |  | 1.03 (0.87-1.22) | 0.7154 | 1.04 (0.79-1.37) | 0.7761 |
| Model 2, HR (95%CI) | 1.00 (0.78-1.28) | 0.72 (0.53-0.97) | 1.03 (0.78-1.35) | 1.00 (0.72-1.39) |  | 1.03 (0.87-1.21) | 0.7470 | 1.03 (0.79-1.36) | 0.8080 |
| Lung (C34) |  |  |  |  |  |  |  |  |  |
| Cases | 243 | 172 | 137 | 104 |  | 656 | | | |
| Model 0, HR (95% CI) | 1.00 (0.88-1.14) | 0.84 (0.73-0.98) | 0.79 (0.67-0.94) | 0.81 (0.66-0.98) |  | **0.91 (0.83-0.99)** | **0.0361** | **0.85 (0.73-0.99)** | **0.0377** |
| Model 1, HR (95% CI) | 1.00 (0.88-1.14) | 0.89 (0.76-1.03) | 0.86 (0.72-1.01) | 0.86 (0.71-1.05) |  | 0.94 (0.85-1.03) | 0.1613 | 0.90 (0.77-1.05) | 0.1641 |
| Model 2, HR (95%CI) | 1.00 (0.88-1.14) | 0.92 (0.79-1.06) | 0.90 (0.76-1.06) | 0.92 (0.75-1.12) |  | 0.96 (0.88-1.05) | 0.3982 | 0.94 (0.80-1.09) | 0.4026 |
| Melanoma (C43) |  |  |  |  |  |  |  |  |  |
| Cases | 109 | 109 | 143 | 108 |  | 469 | | | |
| Model 0, HR (95% CI) | 1.00 (0.83-1.21) | 1.11 (0.92-1.33) | 1.62 (1.37-1.90) | 1.48 (1.22-1.80) |  | **1.20 (1.08-1.33)** | **0.0005** | **1.34 (1.13-1.59)** | **0.0009** |
| Model 1, HR (95% CI) | 1.00 (0.82-1.21) | 1.11 (0.92-1.34) | 1.63 (1.38-1.92) | 1.51 (1.24-1.84) |  | **1.21 (1.09-1.34)** | **0.0004** | **1.35 (1.14-1.61)** | **0.0006** |
| Model 2, HR (95%CI) | 1.00 (0.82-1.22) | 1.10 (0.91-1.33) | 1.61 (1.37-1.90) | 1.49 (1.23-1.82) |  | **1.21 (1.09-1.34)** | **0.0005** | **1.34 (1.13-1.60)** | **0.0009** |
| Meothelioma (C45) |  |  |  |  |  |  |  |  |  |
| Cases | 30 | 38 | 23 | 21 |  | 112 | | | |
| Model 0, HR (95% CI) | 1.00 (0.70-1.44) | 1.56 (1.14-2.14) | 1.18 (0.78-1.77) | 1.59 (1.03-2.46) |  | 1.15 (0.93-1.43) | 0.1915 | 1.29 (0.90-1.84) | 0.1665 |
| Model 1, HR (95% CI) | 1.00 (0.69-1.44) | 1.58 (1.15-2.17) | 1.20 (0.80-1.80) | 1.61 (1.04-2.49) |  | 1.16 (0.93-1.44) | 0.1795 | 1.30 (0.91-1.85) | 0.1557 |
| Model 2, HR (95%CI) | 1.00 (0.69-1.44) | 1.56 (1.13-2.13) | 1.17 (0.78-1.76) | 1.56 (1.01-2.42) |  | 1.15 (0.92-1.42) | 0.2206 | 1.27 (0.89-1.82) | 0.1925 |
| Kidney (C64-65) |  |  |  |  |  |  |  |  |  |
| Cases | 93 | 94 | 63 | 48 |  | 298 | | | |
| Model 0, HR (95% CI) | 1.00 (0.81-1.23) | 1.13 (0.92-1.38) | 0.86 (0.67-1.10) | 0.81 (0.61-1.09) |  | 0.90 (0.79-1.04) | 0.1478 | 0.85 (0.68-1.07) | 0.1670 |
| Model 1, HR (95% CI) | 1.00 (0.81-1.23) | 1.19 (0.98-1.46) | 0.95 (0.74-1.22) | 0.92 (0.69-1.23) |  | 0.95 (0.83-1.09) | 0.4771 | 0.93 (0.74-1.16) | 0.5153 |
| Model 2, HR (95%CI) | 1.00 (0.81-1.24) | 1.19 (0.97-1.45) | 0.94 (0.73-1.20) | 0.90 (0.67-1.20) |  | 0.94 (0.82-1.08) | 0.3935 | 0.91 (0.73-1.15) | 0.4282 |
| Bladder (C67) |  |  |  |  |  |  |  |  |  |
| Cases | 94 | 65 | 65 | 35 |  | 259 | | | |
| Model 0, HR (95% CI) | 1.00 (0.81-1.23) | 0.82 (0.64-1.05) | 1.00 (0.78-1.27) | 0.76 (0.54-1.06) |  | 0.92 (0.80-1.07) | 0.2939 | 0.87 (0.68-1.11) | 0.2579 |
| Model 1, HR (95% CI) | 1.00 (0.81-1.23) | 0.85 (0.67-1.08) | 1.06 (0.83-1.35) | 0.82 (0.59-1.15) |  | 0.96 (0.82-1.11) | 0.5593 | 0.92 (0.72-1.18) | 0.5034 |
| Model 2, HR (95%CI) | 1.00 (0.81-1.23) | 0.85 (0.67-1.08) | 1.05 (0.82-1.34) | 0.81 (0.58-1.14) |  | 0.95 (0.82-1.11) | 0.5282 | 0.91 (0.71-1.17) | 0.4742 |
| Prostate (C60) |  |  |  |  |  |  |  |  |  |
| Cases | 1043 | 1006 | 840 | 661 |  | 3550 | | | |
| Model 0, HR (95% CI) | 1.00 (0.94-1.06) | 1.12 (1.06-1.19) | 1.12 (1.04-1.20) | 1.20 (1.11-1.30) |  | **1.07 (1.03-1.11)** | **0.0004** | **1.12 (1.05-1.20)** | **0.0003** |
| Model 1, HR (95% CI) | 1.00 (0.94-1.06) | 1.11 (1.04-1.18) | 1.10 (1.02-1.17) | 1.17 (1.08-1.27) |  | **1.06 (1.02-1.10)** | **0.0029** | **1.10 (1.04-1.18)** | **0.0024** |
| Model 2, HR (95%CI) | 1.00 (0.94-1.06) | 1.10 (1.03-1.17) | 1.08 (1.01-1.15) | 1.15 (1.06-1.24) |  | **1.05 (1.01-1.09)** | **0.0103** | **1.09 (1.02-1.16)** | **0.0088** |
| Brain (C71) |  |  |  |  |  |  |  |  |  |
| Cases | 44 | 45 | 42 | 32 |  | 163 | | | |
| Model 0, HR (95% CI) | 1.00 (0.74-1.35) | 1.10 (0.82-1.47) | 1.13 (0.84-1.53) | 1.03 (0.72-1.47) |  | 1.02 (0.85-1.21) | 0.8691 | 1.02 (0.76-1.38) | 0.8820 |
| Model 1, HR (95% CI) | 1.00 (0.74-1.36) | 1.09 (0.82-1.46) | 1.12 (0.83-1.52) | 1.02 (0.71-1.46) |  | 1.01 (0.85-1.21) | 0.8930 | 1.02 (0.75-1.37) | 0.9043 |
| Model 2, HR (95%CI) | 1.00 (0.74-1.36) | 1.10 (0.82-1.48) | 1.12 (0.83-1.52) | 1.02 (0.71-1.46) |  | 1.01 (0.84-1.21) | 0.9068 | 1.02 (0.75-1.37) | 0.9171 |
| Non-Hodgkin Lymphoma (C82-85) | |  |  |  |  |  |  |  |  |
| Cases | 118 | 97 | 91 | 62 |  | 368 | | | |
| Model 0, HR (95% CI) | 1.00 (0.83-1.20) | 0.93 (0.76-1.14) | 0.99 (0.81-1.22) | 0.84 (0.65-1.09) |  | 0.95 (0.84-1.07) | 0.3730 | 0.91 (0.74-1.11) | 0.3501 |
| Model 1, HR (95% CI) | 1.00 (0.83-1.21) | 0.93 (0.76-1.13) | 1.00 (0.81-1.22) | 0.85 (0.66-1.10) |  | 0.95 (0.84-1.07) | 0.4127 | 0.91 (0.75-1.12) | 0.3882 |
| Model 2, HR (95%CI) | 1.00 (0.83-1.21) | 0.93 (0.76-1.13) | 0.99 (0.80-1.21) | 0.84 (0.65-1.08) |  | 0.94 (0.84-1.07) | 0.3656 | 0.91 (0.74-1.11) | 0.3432 |
| Diffuse non-Hodgkins lymphoma (C83) | |  |  |  |  |  |  |  |  |
| Cases | 69 | 53 | 55 | 26 |  | 203 | | | |
| Model 0, HR (95% CI) | 1.00 (0.79-1.27) | 0.89 (0.68-1.16) | 1.07 (0.82-1.40) | 0.66 (0.45-0.98) |  | 0.90 (0.76-1.06) | 0.1920 | 0.82 (0.62-1.08) | 0.1629 |
| Model 1, HR (95% CI) | 1.00 (0.78-1.28) | 0.90 (0.69-1.18) | 1.09 (0.84-1.42) | 0.68 (0.46-1.01) |  | 0.91 (0.77-1.07) | 0.2571 | 0.84 (0.63-1.11) | 0.2207 |
| Model 2, HR (95%CI) | 1.00 (0.78-1.28) | 0.90 (0.69-1.17) | 1.08 (0.83-1.40) | 0.67 (0.45-1.00) |  | 0.90 (0.76-1.07) | 0.2226 | 0.83 (0.63-1.10) | 0.1901 |
| Multiple Myeloma (C90) | |  |  |  |  |  |  |  |  |
| Cases | 42 | 35 | 34 | 36 |  | 147 | | | |
| Model 0, HR (95% CI) | 1.00 (0.73-1.36) | 0.97 (0.70-1.35) | 1.09 (0.78-1.53) | 1.46 (1.04-2.06) |  | 1.17 (0.97-1.41) | 0.0996 | 1.30 (0.95-1.77) | 0.0975 |
| Model 1, HR (95% CI) | 1.00 (0.73-1.37) | 0.99 (0.71-1.37) | 1.12 (0.80-1.56) | 1.52 (1.08-2.14) |  | 1.19 (0.98-1.43) | 0.0730 | 1.33 (0.98-1.82) | 0.0711 |
| Model 2, HR (95%CI) | 1.00 (0.73-1.37) | 0.99 (0.71-1.37) | 1.11 (0.79-1.55) | 1.50 (1.06-2.12) |  | 1.18 (0.98-1.43) | 0.0833 | 1.32 (0.97-1.81) | 0.0813 |
| Leukemia (C91-95) |  |  |  |  |  |  |  |  |  |
| Cases | 101 | 70 | 51 | 44 |  | 266 | | | |
| Model 0, HR (95% CI) | 1.00 (0.82-1.22) | 0.79 (0.63-1.00) | 0.67 (0.51-0.88) | 0.73 (0.54-0.99) |  | **0.86 (0.74-0.99)** | **0.0385** | **0.78 (0.61-0.99)** | **0.0426** |
| Model 1, HR (95% CI) | 1.00 (0.82-1.23) | 0.79 (0.63-1.00) | 0.66 (0.50-0.87) | 0.72 (0.53-0.98) |  | **0.85 (0.74-0.99)** | **0.0347** | **0.77 (0.60-0.99)** | **0.0386** |
| Model 2, HR (95%CI) | 1.00 (0.81-1.23) | 0.80 (0.63-1.01) | 0.67 (0.51-0.88) | 0.73 (0.54-1.00) |  | **0.86 (0.74-1.00)** | **0.0443** | **0.78 (0.61-1.00)** | **0.0490** |

Estimates bold if associations are statistically significant (p<0.05).

Model 0: Stratified by age group, geographical region and adjusted for Townsend deprivation score

Model 1: Model 0 + age group, geographical region and adjusted for Townsend deprivation score, racial/ethnic group, height, lives with a spouse or partner, body mass index, cigarette smoking, alcohol consumption, and total physical activity.

Model 2: Model 1 + additionally adjusted for serum concentrations of insulin-like growth factor-I, C-reactive protein, glycated hemoglobin (fourths, unknown).

Abbreviations: CI=Confidence interval; HR=hazard ratio; IQR=interquartile range.

Supplementary Table S2: HR and 95% CIs for cancer diagnosis by serum total testosterone concentrations in men

|  | Fourths of total testosterone | | | |  | per 5 nmol/L increment | | | |
| --- | --- | --- | --- | --- | --- | --- | --- | --- | --- |
|  |  |  |  |  |  | Not corrected for regression dilution bias | | Corrected for regression dilution bias | |
|  | 1 | 2 | 3 | 4 |  |  | P_trend_ |  | P_trend_ |
| Whole cohort, n | 45665 | 45642 | 45657 | 45644 |  | 182608 | | | |
| Median (IQR), nmol/L | 8.2 (1.8) | 10.7 (1.1) | 12.9 (1.2) | 16.2 (2.9) |  |  |  |  |  |
| Oral (C00-14) |  |  |  |  |  |  |  |  |  |
| Cases | 67 | 65 | 60 | 53 |  | 245 | | | |
| Model 0, HR (95% CI) | 1.00 (0.79-1.27) | 0.98 (0.77-1.26) | 0.91 (0.70-1.17) | 0.80 (0.61-1.05) |  | 0.87 (0.70-1.08) | 0.1966 | 0.83 (0.63-1.10) | 0.1981 |
| Model 1, HR (95% CI) | 1.00 (0.78-1.28) | 0.98 (0.77-1.25) | 0.89 (0.69-1.14) | 0.72 (0.54-0.95) |  | 0.81 (0.65-1.02) | 0.0696 | 0.76 (0.57-1.02) | 0.0699 |
| Model 2, HR (95%CI) | 1.00 (0.78-1.29) | 1.00 (0.79-1.27) | 0.91 (0.71-1.17) | 0.74 (0.56-0.98) |  | 0.83 (0.66-1.04) | 0.1034 | 0.78 (0.58-1.05) | 0.1037 |
| Esophagus (C15) |  |  |  |  |  |  |  |  |  |
| Cases | 64 | 69 | 50 | 57 |  | 240 | | | |
| Model 0, HR (95% CI) | 1.00 (0.78-1.28) | 1.12 (0.89-1.42) | 0.82 (0.62-1.08) | 0.97 (0.75-1.26) |  | 0.94 (0.75-1.16) | 0.5501 | 0.93 (0.70-1.23) | 0.6003 |
| Model 1, HR (95% CI) | 1.00 (0.78-1.29) | 1.22 (0.97-1.55) | 0.94 (0.71-1.24) | 1.17 (0.89-1.53) |  | 1.05 (0.84-1.32) | 0.6681 | 1.08 (0.80-1.45) | 0.6160 |
| Model 2, HR (95%CI) | 1.00 (0.77-1.29) | 1.26 (0.99-1.59) | 0.98 (0.74-1.29) | 1.25 (0.95-1.63) |  | 1.09 (0.87-1.37) | 0.4456 | 1.13 (0.84-1.53) | 0.4031 |
| Adenocarcinoma, esophagus (C15) | |  |  |  |  |  |  |  |  |
| Cases | 57 | 53 | 42 | 40 |  | 192 | | | |
| Model 0, HR (95% CI) | 1.00 (0.77-1.30) | 0.96 (0.74-1.26) | 0.77 (0.57-1.04) | 0.77 (0.57-1.05) |  | 0.83 (0.65-1.06) | 0.1374 | 0.79 (0.57-1.09) | 0.1468 |
| Model 1, HR (95% CI) | 1.00 (0.76-1.31) | 1.08 (0.83-1.41) | 0.92 (0.68-1.24) | 0.97 (0.71-1.34) |  | 0.96 (0.74-1.24) | 0.7516 | 0.95 (0.68-1.33) | 0.7767 |
| Model 2, HR (95%CI) | 1.00 (0.76-1.32) | 1.11 (0.85-1.45) | 0.96 (0.71-1.30) | 1.04 (0.75-1.44) |  | 1.00 (0.77-1.29) | 0.9986 | 1.01 (0.72-1.41) | 0.9728 |
| Stomach (C15) |  |  |  |  |  |  |  |  |  |
| Cases | 41 | 34 | 40 | 37 |  | 152 | | | |
| Model 0, HR (95% CI) | 1.00 (0.74-1.36) | 0.86 (0.61-1.20) | 1.01 (0.74-1.38) | 0.97 (0.70-1.34) |  | 1.01 (0.77-1.32) | 0.9554 | 1.01 (0.71-1.43) | 0.9777 |
| Model 1, HR (95% CI) | 1.00 (0.73-1.38) | 0.91 (0.65-1.28) | 1.11 (0.82-1.52) | 1.09 (0.78-1.52) |  | 1.09 (0.82-1.44) | 0.5646 | 1.11 (0.77-1.60) | 0.5864 |
| Model 2, HR (95%CI) | 1.00 (0.72-1.38) | 0.93 (0.67-1.31) | 1.15 (0.85-1.57) | 1.14 (0.81-1.60) |  | 1.12 (0.84-1.49) | 0.4508 | 1.15 (0.79-1.67) | 0.4709 |
| Colorectum (C18-20) |  |  |  |  |  |  |  |  |  |
| Cases | 319 | 296 | 277 | 232 |  | 1124 | | | |
| Model 0, HR (95% CI) | 1.00 (0.90-1.12) | 0.95 (0.85-1.07) | 0.90 (0.80-1.01) | 0.79 (0.70-0.90) |  | **0.87 (0.78-0.96)** | **0.0055** | **0.83 (0.73-0.95)** | **0.0054** |
| Model 1, HR (95% CI) | 1.00 (0.89-1.12) | 0.97 (0.87-1.09) | 0.94 (0.84-1.06) | 0.85 (0.74-0.97) |  | 0.90 (0.81-1.00) | 0.0604 | 0.88 (0.76-1.01) | 0.0594 |
| Model 2, HR (95%CI) | 1.00 (0.89-1.12) | 0.98 (0.88-1.10) | 0.96 (0.85-1.08) | 0.87 (0.77-1.00) |  | 0.92 (0.83-1.03) | 0.1353 | 0.90 (0.78-1.03) | 0.1329 |
| Colon (C18) |  |  |  |  |  |  |  |  |  |
| Cases | 202 | 184 | 170 | 137 |  | 693 | | | |
| Model 0, HR (95% CI) | 1.00 (0.87-1.15) | 0.93 (0.81-1.08) | 0.87 (0.75-1.02) | 0.74 (0.62-0.87) |  | **0.83 (0.73-0.95)** | **0.0051** | **0.79 (0.66-0.93)** | **0.0050** |
| Model 1, HR (95% CI) | 1.00 (0.87-1.15) | 0.98 (0.84-1.13) | 0.94 (0.81-1.10) | 0.83 (0.70-0.98) |  | 0.89 (0.78-1.02) | 0.0992 | 0.86 (0.72-1.03) | 0.0974 |
| Model 2, HR (95%CI) | 1.00 (0.87-1.16) | 0.98 (0.85-1.13) | 0.96 (0.82-1.11) | 0.85 (0.72-1.01) |  | 0.91 (0.79-1.04) | 0.1637 | 0.88 (0.74-1.05) | 0.1607 |
| Rectum (C19-20) |  |  |  |  |  |  |  |  |  |
| Cases | 118 | 115 | 108 | 95 |  | 436 | | | |
| Model 0, HR (95% CI) | 1.00 (0.83-1.20) | 1.00 (0.83-1.20) | 0.95 (0.79-1.15) | 0.87 (0.72-1.07) |  | 0.92 (0.78-1.08) | 0.2945 | 0.89 (0.72-1.10) | 0.2962 |
| Model 1, HR (95% CI) | 1.00 (0.83-1.20) | 0.99 (0.82-1.19) | 0.94 (0.78-1.14) | 0.87 (0.71-1.07) |  | 0.91 (0.77-1.08) | 0.2974 | 0.89 (0.72-1.11) | 0.2984 |
| Model 2, HR (95%CI) | 1.00 (0.83-1.21) | 1.01 (0.84-1.21) | 0.97 (0.80-1.17) | 0.91 (0.74-1.12) |  | 0.94 (0.79-1.11) | 0.4618 | 0.92 (0.74-1.15) | 0.4628 |
| Liver (C22) |  |  |  |  |  |  |  |  |  |
| Cases | 22 | 19 | 18 | 41 |  | 100 | | | |
| Model 0, HR (95% CI) | 1.00 (0.66-1.52) | 0.89 (0.57-1.40) | 0.85 (0.54-1.36) | 2.02 (1.49-2.75) |  | **1.66 (1.19-2.32)** | **0.0028** | **1.96 (1.27-3.02)** | **0.0023** |
| Model 1, HR (95% CI) | 1.00 (0.65-1.55) | 1.03 (0.66-1.62) | 1.07 (0.68-1.69) | 2.70 (1.95-3.74) |  | **1.98 (1.40-2.80)** | **0.0001** | **2.45 (1.56-3.84)** | **0.0001** |
| Model 2, HR (95%CI) | 1.00 (0.64-1.56) | 1.06 (0.68-1.66) | 1.09 (0.69-1.72) | 2.62 (1.88-3.64) |  | **1.93 (1.36-2.73)** | **0.0002** | **2.36 (1.50-3.71)** | **0.0002** |
| Pancreas (C25) |  |  |  |  |  |  |  |  |  |
| Cases | 64 | 45 | 52 | 52 |  | 213 | | | |
| Model 0, HR (95% CI) | 1.00 (0.78-1.28) | 0.73 (0.54-0.98) | 0.85 (0.65-1.12) | 0.90 (0.68-1.18) |  | 0.96 (0.76-1.21) | 0.7436 | 0.95 (0.70-1.28) | 0.7237 |
| Model 1, HR (95% CI) | 1.00 (0.77-1.29) | 0.75 (0.56-1.00) | 0.89 (0.68-1.16) | 0.94 (0.71-1.24) |  | 0.99 (0.78-1.26) | 0.9336 | 0.98 (0.72-1.34) | 0.9107 |
| Model 2, HR (95%CI) | 1.00 (0.77-1.30) | 0.77 (0.58-1.04) | 0.93 (0.71-1.22) | 1.02 (0.77-1.35) |  | 1.04 (0.82-1.33) | 0.7384 | 1.05 (0.77-1.44) | 0.7599 |
| Lung (C34) |  |  |  |  |  |  |  |  |  |
| Cases | 190 | 156 | 179 | 180 |  | 705 | | | |
| Model 0, HR (95% CI) | 1.00 (0.87-1.15) | 0.86 (0.73-1.01) | 0.99 (0.86-1.15) | 1.02 (0.88-1.18) |  | 1.04 (0.92-1.18) | 0.5487 | 1.05 (0.89-1.23) | 0.5770 |
| Model 1, HR (95% CI) | 1.00 (0.86-1.16) | 0.83 (0.71-0.98) | 0.92 (0.79-1.06) | 0.87 (0.75-1.01) |  | 0.94 (0.82-1.07) | 0.3396 | 0.92 (0.77-1.09) | 0.3179 |
| Model 2, HR (95%CI) | 1.00 (0.86-1.16) | 0.87 (0.74-1.01) | 0.98 (0.85-1.14) | 0.95 (0.81-1.11) |  | 0.99 (0.87-1.13) | 0.9027 | 0.99 (0.83-1.17) | 0.8647 |
| Melanoma (C43) |  |  |  |  |  |  |  |  |  |
| Cases | 124 | 127 | 131 | 140 |  | 522 | | | |
| Model 0, HR (95% CI) | 1.00 (0.84-1.19) | 1.04 (0.88-1.24) | 1.09 (0.92-1.29) | 1.23 (1.04-1.45) |  | 1.14 (0.98-1.32) | 0.0797 | 1.19 (0.98-1.43) | 0.0785 |
| Model 1, HR (95% CI) | 1.00 (0.83-1.20) | 1.07 (0.90-1.27) | 1.15 (0.97-1.37) | 1.35 (1.14-1.60) |  | **1.21 (1.04-1.40)** | **0.0152** | **1.28 (1.05-1.55)** | **0.0150** |
| Model 2, HR (95%CI) | 1.00 (0.83-1.20) | 1.06 (0.89-1.26) | 1.13 (0.96-1.34) | 1.32 (1.11-1.57) |  | **1.19 (1.02-1.39)** | **0.0243** | **1.26 (1.03-1.53)** | **0.0240** |
| Meothelioma (C45) |  |  |  |  |  |  |  |  |  |
| Cases | 24 | 21 | 47 | 34 |  | 126 | | | |
| Model 0, HR (95% CI) | 1.00 (0.67-1.49) | 0.92 (0.60-1.40) | 2.10 (1.58-2.80) | 1.63 (1.16-2.28) |  | **1.47 (1.10-1.98)** | **0.0102** | **1.61 (1.10-2.37)** | **0.0148** |
| Model 1, HR (95% CI) | 1.00 (0.66-1.51) | 0.89 (0.58-1.36) | 2.11 (1.59-2.80) | 1.64 (1.16-2.31) |  | **1.49 (1.10-2.02)** | **0.0106** | **1.63 (1.10-2.42)** | **0.0157** |
| Model 2, HR (95%CI) | 1.00 (0.66-1.51) | 0.86 (0.56-1.32) | 2.06 (1.55-2.73) | 1.57 (1.11-2.22) |  | **1.46 (1.07-1.99)** | **0.0166** | **1.59 (1.06-2.37)** | **0.0245** |
| Prostate (C60) |  |  |  |  |  |  |  |  |  |
| Cases | 988 | 972 | 981 | 904 |  | 3845 | | | |
| Model 0, HR (95% CI) | 1.00 (0.94-1.06) | 1.01 (0.95-1.08) | 1.04 (0.98-1.11) | 1.02 (0.95-1.09) |  | 1.01 (0.96-1.07) | 0.6263 | 1.02 (0.95-1.09) | 0.6433 |
| Model 1, HR (95% CI) | 1.00 (0.94-1.07) | 0.99 (0.93-1.06) | 1.01 (0.95-1.07) | 0.98 (0.92-1.05) |  | 0.99 (0.94-1.05) | 0.7114 | 0.99 (0.92-1.06) | 0.6957 |
| Model 2, HR (95%CI) | 1.00 (0.94-1.07) | 0.99 (0.93-1.05) | 1.00 (0.94-1.07) | 0.98 (0.91-1.05) |  | 0.99 (0.93-1.05) | 0.7144 | 0.99 (0.92-1.06) | 0.6984 |
| Kidney (C64-65) |  |  |  |  |  |  |  |  |  |
| Cases | 103 | 86 | 67 | 73 |  | 329 | | | |
| Model 0, HR (95% CI) | 1.00 (0.82-1.21) | 0.86 (0.70-1.06) | 0.68 (0.53-0.86) | 0.77 (0.62-0.97) |  | **0.83 (0.68-1.00)** | **0.0467** | 0.79 (0.61-1.00) | 0.0530 |
| Model 1, HR (95% CI) | 1.00 (0.82-1.22) | 0.94 (0.76-1.15) | 0.77 (0.61-0.98) | 0.93 (0.73-1.17) |  | 0.93 (0.76-1.13) | 0.4419 | 0.91 (0.71-1.18) | 0.4743 |
| Model 2, HR (95%CI) | 1.00 (0.82-1.23) | 0.95 (0.77-1.17) | 0.79 (0.62-1.00) | 0.96 (0.76-1.22) |  | 0.95 (0.78-1.16) | 0.6018 | 0.94 (0.73-1.22) | 0.6401 |
| Bladder (C67) |  |  |  |  |  |  |  |  |  |
| Cases | 88 | 58 | 73 | 66 |  | 285 | | | |
| Model 0, HR (95% CI) | 1.00 (0.81-1.23) | 0.68 (0.53-0.88) | 0.87 (0.69-1.10) | 0.84 (0.66-1.07) |  | 0.94 (0.77-1.14) | 0.5236 | 0.91 (0.70-1.18) | 0.4865 |
| Model 1, HR (95% CI) | 1.00 (0.80-1.24) | 0.71 (0.55-0.91) | 0.94 (0.74-1.18) | 0.93 (0.73-1.20) |  | 1.00 (0.81-1.23) | 0.9991 | 0.99 (0.76-1.30) | 0.9495 |
| Model 2, HR (95%CI) | 1.00 (0.80-1.25) | 0.71 (0.55-0.92) | 0.95 (0.75-1.19) | 0.95 (0.74-1.22) |  | 1.01 (0.82-1.25) | 0.9269 | 1.00 (0.76-1.32) | 0.9781 |
| Brain (C71) |  |  |  |  |  |  |  |  |  |
| Cases | 58 | 48 | 39 | 44 |  | 189 | | | |
| Model 0, HR (95% CI) | 1.00 (0.77-1.29) | 0.85 (0.64-1.13) | 0.69 (0.50-0.94) | 0.81 (0.60-1.09) |  | 0.85 (0.67-1.09) | 0.2094 | 0.82 (0.59-1.13) | 0.2242 |
| Model 1, HR (95% CI) | 1.00 (0.76-1.31) | 0.85 (0.64-1.12) | 0.67 (0.49-0.91) | 0.76 (0.56-1.04) |  | 0.82 (0.64-1.07) | 0.1398 | 0.78 (0.56-1.09) | 0.1517 |
| Model 2, HR (95%CI) | 1.00 (0.76-1.31) | 0.85 (0.64-1.13) | 0.67 (0.49-0.92) | 0.77 (0.57-1.05) |  | 0.83 (0.64-1.08) | 0.1629 | 0.79 (0.56-1.11) | 0.1763 |
| Non-Hodgkin Lymphoma (C82-85) | |  |  |  |  |  |  |  |  |
| Cases | 124 | 97 | 88 | 99 |  | 408 | | | |
| Model 0, HR (95% CI) | 1.00 (0.84-1.19) | 0.80 (0.66-0.98) | 0.73 (0.60-0.91) | 0.87 (0.71-1.06) |  | 0.91 (0.77-1.08) | 0.2749 | 0.89 (0.71-1.11) | 0.2884 |
| Model 1, HR (95% CI) | 1.00 (0.83-1.20) | 0.80 (0.66-0.97) | 0.73 (0.59-0.90) | 0.87 (0.71-1.07) |  | 0.91 (0.77-1.09) | 0.3074 | 0.89 (0.71-1.12) | 0.3228 |
| Model 2, HR (95%CI) | 1.00 (0.83-1.20) | 0.80 (0.66-0.98) | 0.74 (0.60-0.91) | 0.89 (0.72-1.09) |  | 0.92 (0.78-1.10) | 0.3827 | 0.91 (0.72-1.14) | 0.4004 |
| Diffuse non-Hodgkins lymphoma (C83) | |  |  |  |  |  |  |  |  |
| Cases | 70 | 52 | 48 | 54 |  | 224 | | | |
| Model 0, HR (95% CI) | 1.00 (0.79-1.26) | 0.76 (0.58-1.00) | 0.71 (0.54-0.95) | 0.85 (0.65-1.11) |  | 0.90 (0.72-1.13) | 0.3583 | 0.87 (0.65-1.17) | 0.3684 |
| Model 1, HR (95% CI) | 1.00 (0.78-1.28) | 0.77 (0.59-1.01) | 0.73 (0.55-0.97) | 0.88 (0.67-1.16) |  | 0.92 (0.73-1.16) | 0.4904 | 0.90 (0.66-1.22) | 0.5026 |
| Model 2, HR (95%CI) | 1.00 (0.78-1.28) | 0.77 (0.59-1.01) | 0.73 (0.55-0.97) | 0.90 (0.68-1.18) |  | 0.93 (0.73-1.18) | 0.5596 | 0.91 (0.67-1.25) | 0.5722 |
| Multiple Myeloma (C90) | |  |  |  |  |  |  |  |  |
| Cases | 51 | 36 | 37 | 39 |  | 163 | | | |
| Model 0, HR (95% CI) | 1.00 (0.76-1.32) | 0.72 (0.52-1.00) | 0.76 (0.55-1.04) | 0.84 (0.61-1.15) |  | 0.90 (0.69-1.18) | 0.4583 | 0.88 (0.62-1.24) | 0.4553 |
| Model 1, HR (95% CI) | 1.00 (0.75-1.33) | 0.72 (0.52-1.00) | 0.76 (0.55-1.05) | 0.83 (0.60-1.15) |  | 0.90 (0.69-1.19) | 0.4764 | 0.88 (0.61-1.26) | 0.4728 |
| Model 2, HR (95%CI) | 1.00 (0.75-1.34) | 0.72 (0.52-1.00) | 0.77 (0.56-1.06) | 0.85 (0.62-1.18) |  | 0.92 (0.70-1.21) | 0.5538 | 0.90 (0.62-1.29) | 0.5492 |
| Leukemia (C91-95) |  |  |  |  |  |  |  |  |  |
| Cases | 90 | 66 | 65 | 64 |  | 285 | | | |
| Model 0, HR (95% CI) | 1.00 (0.81-1.23) | 0.76 (0.59-0.96) | 0.75 (0.59-0.96) | 0.78 (0.61-1.00) |  | 0.86 (0.70-1.05) | 0.1472 | 0.82 (0.63-1.07) | 0.1463 |
| Model 1, HR (95% CI) | 1.00 (0.81-1.24) | 0.73 (0.58-0.93) | 0.72 (0.56-0.91) | 0.73 (0.56-0.93) |  | 0.82 (0.67-1.01) | 0.0685 | 0.78 (0.59-1.02) | 0.0681 |
| Model 2, HR (95%CI) | 1.00 (0.80-1.24) | 0.74 (0.58-0.94) | 0.72 (0.56-0.92) | 0.72 (0.56-0.93) |  | 0.82 (0.67-1.02) | 0.0690 | 0.77 (0.59-1.02) | 0.0687 |

Estimates bold if associations are statistically significant (p<0.05).

Model 0: Stratified by age group, geographical region and adjusted for Townsend deprivation score

Model 1: Model 0 + age group, geographical region and adjusted for Townsend deprivation score, racial/ethnic group, height, lives with a spouse or partner, body mass index, cigarette smoking, alcohol consumption, and total physical activity.

Model 2: Model 1 + additionally adjusted for serum concentrations of insulin-like growth factor-I, C-reactive protein, glycated hemoglobin (fourths, unknown).

Abbreviations: CI=Confidence interval; HR=hazard ratio; IQR=interquartile range.

Supplementary Table S3: HR and 95% CIs for cancer diagnosis by serum SHBG concentrations in men

|  | Fourths of SHBG | | | |  | per 10 nmol/L increment | | | |
| --- | --- | --- | --- | --- | --- | --- | --- | --- | --- |
|  |  |  |  |  |  | Not corrected for regression dilution bias | | Corrected for regression dilution bias | |
|  | 1 | 2 | 3 | 4 |  |  | P_trend_ |  | P_trend_ |
| Whole cohort, n | 42220 | 42220 | 42197 | 42198 |  | 168835 | | |  |
| Median (IQR), nmol/L | 22.8 (6.6) | 32.6 (4.4) | 42.0 (5.4) | 57.8 (14.6) |  |  |  |  |  |
| Oral (C00-14) |  |  |  |  |  |  |  |  |  |
| Cases | 47 | 58 | 60 | 65 |  | 230 | | | |
| Model 0, HR (95% CI) | 1.00 (0.75-1.34) | 1.18 (0.91-1.52) | 1.16 (0.91-1.50) | 1.22 (0.95-1.56) |  | 1.05 (0.94-1.16) | 0.3946 | 1.05 (0.94-1.16) | 0.3907 |
| Model 1, HR (95% CI) | 1.00 (0.74-1.36) | 1.13 (0.88-1.47) | 1.08 (0.84-1.39) | 1.03 (0.79-1.34) |  | 1.00 (0.89-1.11) | 0.9374 | 1.00 (0.89-1.11) | 0.9462 |
| Model 2, HR (95%CI) | 1.00 (0.73-1.36) | 1.15 (0.89-1.49) | 1.10 (0.86-1.41) | 1.03 (0.79-1.36) |  | 1.00 (0.89-1.12) | 0.9579 | 1.00 (0.89-1.12) | 0.9685 |
| Esophagus (C15) |  |  |  |  |  |  |  |  |  |
| Cases | 43 | 45 | 58 | 72 |  | 218 | | | |
| Model 0, HR (95% CI) | 1.00 (0.74-1.36) | 0.81 (0.60-1.08) | 0.89 (0.69-1.15) | 0.94 (0.74-1.19) |  | 1.00 (0.90-1.12) | 0.9339 | 1.00 (0.90-1.12) | 0.9443 |
| Model 1, HR (95% CI) | 1.00 (0.73-1.37) | 0.85 (0.63-1.13) | 0.96 (0.75-1.24) | 1.07 (0.84-1.37) |  | 1.04 (0.93-1.16) | 0.4655 | 1.04 (0.93-1.17) | 0.4733 |
| Model 2, HR (95%CI) | 1.00 (0.73-1.38) | 0.87 (0.65-1.17) | 1.02 (0.79-1.31) | 1.17 (0.91-1.51) |  | 1.07 (0.95-1.20) | 0.2487 | 1.07 (0.95-1.20) | 0.2539 |
| Adenocarcinoma, esophagus (C15) | |  |  |  |  |  |  |  |  |
| Cases | 34 | 35 | 48 | 58 |  | 175 | | | |
| Model 0, HR (95% CI) | 1.00 (0.71-1.41) | 0.80 (0.57-1.11) | 0.93 (0.70-1.23) | 0.96 (0.74-1.25) |  | 1.01 (0.90-1.14) | 0.8263 | 1.01 (0.90-1.14) | 0.8329 |
| Model 1, HR (95% CI) | 1.00 (0.70-1.42) | 0.85 (0.61-1.19) | 1.05 (0.79-1.38) | 1.16 (0.88-1.53) |  | 1.07 (0.94-1.21) | 0.3013 | 1.07 (0.94-1.21) | 0.3047 |
| Model 2, HR (95%CI) | 1.00 (0.70-1.43) | 0.89 (0.64-1.24) | 1.13 (0.85-1.49) | 1.30 (0.98-1.72) |  | 1.10 (0.97-1.25) | 0.1318 | 1.10 (0.97-1.26) | 0.1335 |
| Stomach (C15) |  |  |  |  |  |  |  |  |  |
| Cases | 20 | 25 | 40 | 58 |  | 143 | | | |
| Model 0, HR (95% CI) | 1.00 (0.64-1.57) | 0.99 (0.67-1.46) | 1.38 (1.02-1.88) | 1.71 (1.31-2.23) |  | **1.19 (1.04-1.36)** | **0.0097** | **1.20 (1.04-1.37)** | **0.0097** |
| Model 1, HR (95% CI) | 1.00 (0.63-1.58) | 1.01 (0.68-1.50) | 1.42 (1.05-1.93) | 1.80 (1.36-2.39) |  | **1.21 (1.05-1.39)** | **0.0076** | **1.21 (1.05-1.40)** | **0.0077** |
| Model 2, HR (95%CI) | 1.00 (0.63-1.59) | 1.04 (0.70-1.55) | 1.49 (1.10-2.02) | 1.94 (1.46-2.59) |  | **1.23 (1.07-1.42)** | **0.0039** | **1.24 (1.07-1.43)** | **0.0040** |
| Colorectum (C18-20) |  |  |  |  |  |  |  |  |  |
| Cases | 237 | 255 | 264 | 291 |  | 1047 | | | |
| Model 0, HR (95% CI) | 1.00 (0.88-1.14) | 0.84 (0.74-0.95) | 0.76 (0.67-0.85) | 0.73 (0.65-0.82) |  | **0.92 (0.87-0.96)** | **0.0006** | **0.92 (0.87-0.96)** | **0.0005** |
| Model 1, HR (95% CI) | 1.00 (0.87-1.14) | 0.86 (0.76-0.97) | 0.79 (0.70-0.89) | 0.79 (0.70-0.89) |  | **0.94 (0.89-0.99)** | **0.0202** | **0.94 (0.89-0.99)** | **0.0184** |
| Model 2, HR (95%CI) | 1.00 (0.87-1.15) | 0.87 (0.77-0.98) | 0.81 (0.72-0.91) | 0.83 (0.73-0.94) |  | 0.95 (0.91-1.01) | 0.0782 | 0.95 (0.90-1.00) | 0.0723 |
| Colon (C18) |  |  |  |  |  |  |  |  |  |
| Cases | 143 | 150 | 173 | 177 |  | 643 | | | |
| Model 0, HR (95% CI) | 1.00 (0.84-1.18) | 0.82 (0.70-0.96) | 0.82 (0.71-0.95) | 0.73 (0.63-0.85) |  | **0.93 (0.87-0.99)** | **0.0158** | **0.92 (0.87-0.99)** | **0.0154** |
| Model 1, HR (95% CI) | 1.00 (0.84-1.19) | 0.85 (0.73-1.00) | 0.89 (0.76-1.03) | 0.85 (0.72-0.99) |  | 0.96 (0.90-1.03) | 0.2752 | 0.96 (0.90-1.03) | 0.2708 |
| Model 2, HR (95%CI) | 1.00 (0.84-1.19) | 0.86 (0.74-1.01) | 0.91 (0.78-1.05) | 0.88 (0.75-1.04) |  | 0.98 (0.91-1.04) | 0.4803 | 0.98 (0.91-1.04) | 0.4739 |
| Rectum (C19-20) |  |  |  |  |  |  |  |  |  |
| Cases | 95 | 107 | 92 | 115 |  | 409 | | | |
| Model 0, HR (95% CI) | 1.00 (0.81-1.23) | 0.88 (0.73-1.07) | 0.66 (0.54-0.81) | 0.72 (0.59-0.86) |  | **0.90 (0.84-0.98)** | **0.0126** | **0.90 (0.83-0.98)** | **0.0112** |
| Model 1, HR (95% CI) | 1.00 (0.81-1.24) | 0.87 (0.72-1.06) | 0.65 (0.53-0.80) | 0.71 (0.59-0.87) |  | **0.90 (0.83-0.98)** | **0.0176** | **0.90 (0.83-0.98)** | **0.0156** |
| Model 2, HR (95%CI) | 1.00 (0.80-1.24) | 0.89 (0.74-1.07) | 0.67 (0.55-0.82) | 0.75 (0.62-0.92) |  | 0.92 (0.84-1.00) | 0.0515 | **0.92 (0.84-1.00)** | **0.0464** |
| Liver (C22) |  |  |  |  |  |  |  |  |  |
| Cases | 14 | 10 | 21 | 52 |  | 97 | | | |
| Model 0, HR (95% CI) | 1.00 (0.58-1.71) | 0.58 (0.31-1.08) | 1.10 (0.72-1.67) | 2.42 (1.83-3.21) |  | **1.45 (1.23-1.72)** | **<0.0001** | **1.46 (1.23-1.73)** | **<0.0001** |
| Model 1, HR (95% CI) | 1.00 (0.58-1.74) | 0.65 (0.35-1.20) | 1.31 (0.86-1.99) | 3.12 (2.30-4.23) |  | **1.55 (1.30-1.84)** | **<0.0001** | **1.56 (1.31-1.87)** | **<0.0001** |
| Model 2, HR (95%CI) | 1.00 (0.57-1.75) | 0.61 (0.33-1.14) | 1.20 (0.79-1.81) | 2.55 (1.86-3.50) |  | **1.46 (1.22-1.74)** | **<0.0001** | **1.47 (1.22-1.76)** | **<0.0001** |
| Pancreas (C25) |  |  |  |  |  |  |  |  |  |
| Cases | 37 | 45 | 57 | 60 |  | 199 | | | |
| Model 0, HR (95% CI) | 1.00 (0.72-1.39) | 0.94 (0.70-1.26) | 1.03 (0.79-1.33) | 0.94 (0.73-1.22) |  | 0.99 (0.88-1.10) | 0.8375 | 0.99 (0.88-1.11) | 0.8431 |
| Model 1, HR (95% CI) | 1.00 (0.71-1.40) | 0.95 (0.71-1.28) | 1.05 (0.81-1.36) | 0.98 (0.74-1.28) |  | 1.00 (0.89-1.12) | 0.9850 | 1.00 (0.89-1.13) | 0.9910 |
| Model 2, HR (95%CI) | 1.00 (0.71-1.41) | 0.99 (0.74-1.33) | 1.13 (0.88-1.46) | 1.11 (0.84-1.46) |  | 1.04 (0.92-1.17) | 0.5692 | 1.04 (0.92-1.17) | 0.5637 |
| Lung (C34) |  |  |  |  |  |  |  |  |  |
| Cases | 91 | 125 | 190 | 253 |  | 659 | | | |
| Model 0, HR (95% CI) | 1.00 (0.81-1.23) | 1.03 (0.87-1.23) | 1.33 (1.15-1.53) | 1.46 (1.29-1.66) |  | **1.12 (1.06-1.19)** | **0.0002** | **1.13 (1.06-1.20)** | **0.0002** |
| Model 1, HR (95% CI) | 1.00 (0.81-1.24) | 0.95 (0.80-1.13) | 1.12 (0.98-1.29) | 1.07 (0.94-1.23) |  | 1.03 (0.96-1.10) | 0.3986 | 1.03 (0.96-1.10) | 0.3890 |
| Model 2, HR (95%CI) | 1.00 (0.81-1.24) | 0.98 (0.82-1.17) | 1.20 (1.04-1.38) | 1.15 (1.00-1.33) |  | 1.05 (0.98-1.12) | 0.1761 | 1.05 (0.98-1.12) | 0.1689 |
| Melanoma (C43) |  |  |  |  |  |  |  |  |  |
| Cases | 98 | 124 | 128 | 122 |  | 472 | | | |
| Model 0, HR (95% CI) | 1.00 (0.82-1.23) | 1.08 (0.90-1.28) | 1.02 (0.86-1.21) | 0.90 (0.75-1.08) |  | 0.96 (0.89-1.03) | 0.2783 | 0.96 (0.89-1.03) | 0.2857 |
| Model 1, HR (95% CI) | 1.00 (0.81-1.23) | 1.10 (0.92-1.31) | 1.06 (0.89-1.26) | 0.98 (0.81-1.19) |  | 0.99 (0.91-1.06) | 0.7147 | 0.99 (0.91-1.07) | 0.7262 |
| Model 2, HR (95%CI) | 1.00 (0.81-1.24) | 1.08 (0.91-1.29) | 1.03 (0.87-1.23) | 0.95 (0.78-1.15) |  | 0.98 (0.90-1.06) | 0.5744 | 0.98 (0.90-1.06) | 0.5843 |
| Meothelioma (C45) |  |  |  |  |  |  |  |  |  |
| Cases | 15 | 23 | 27 | 47 |  | 112 | | | |
| Model 0, HR (95% CI) | 1.00 (0.60-1.67) | 1.08 (0.72-1.62) | 1.03 (0.71-1.50) | 1.46 (1.09-1.96) |  | 1.13 (0.97-1.31) | 0.1153 | 1.13 (0.97-1.32) | 0.1194 |
| Model 1, HR (95% CI) | 1.00 (0.59-1.68) | 1.07 (0.71-1.61) | 1.00 (0.69-1.45) | 1.40 (1.03-1.90) |  | 1.11 (0.95-1.30) | 0.1751 | 1.11 (0.95-1.31) | 0.1810 |
| Model 2, HR (95%CI) | 1.00 (0.59-1.69) | 1.04 (0.69-1.56) | 0.97 (0.67-1.41) | 1.35 (0.99-1.85) |  | 1.11 (0.94-1.30) | 0.2154 | 1.11 (0.94-1.30) | 0.2227 |
| Prostate (C60) |  |  |  |  |  |  |  |  |  |
| Cases | 655 | 908 | 986 | 1026 |  | 3575 | | | |
| Model 0, HR (95% CI) | 1.00 (0.92-1.08) | 1.03 (0.97-1.10) | 0.95 (0.89-1.01) | 0.85 (0.80-0.91) |  | **0.95 (0.92-0.97)** | **0.0001** | **0.95 (0.92-0.97)** | **0.0001** |
| Model 1, HR (95% CI) | 1.00 (0.92-1.08) | 1.01 (0.94-1.08) | 0.92 (0.86-0.98) | 0.81 (0.76-0.87) |  | **0.93 (0.91-0.96)** | **<0.0001** | **0.93 (0.91-0.96)** | **<0.0001** |
| Model 2, HR (95%CI) | 1.00 (0.92-1.08) | 1.01 (0.94-1.07) | 0.92 (0.86-0.97) | 0.81 (0.76-0.87) |  | **0.94 (0.91-0.96)** | **<0.0001** | **0.94 (0.91-0.96)** | **<0.0001** |
| Kidney (C64-65) |  |  |  |  |  |  |  |  |  |
| Cases | 73 | 72 | 76 | 79 |  | 300 | | | |
| Model 0, HR (95% CI) | 1.00 (0.79-1.27) | 0.79 (0.63-0.99) | 0.74 (0.59-0.92) | 0.68 (0.54-0.85) |  | **0.90 (0.82-0.99)** | **0.0287** | **0.90 (0.82-0.99)** | **0.0275** |
| Model 1, HR (95% CI) | 1.00 (0.78-1.28) | 0.84 (0.67-1.05) | 0.82 (0.65-1.02) | 0.79 (0.62-1.00) |  | 0.94 (0.86-1.04) | 0.2190 | 0.94 (0.85-1.04) | 0.2139 |
| Model 2, HR (95%CI) | 1.00 (0.78-1.28) | 0.86 (0.68-1.08) | 0.85 (0.68-1.06) | 0.85 (0.67-1.08) |  | 0.96 (0.87-1.06) | 0.4473 | 0.96 (0.87-1.06) | 0.4392 |
| Bladder (C67) |  |  |  |  |  |  |  |  |  |
| Cases | 36 | 70 | 74 | 80 |  | 260 | | | |
| Model 0, HR (95% CI) | 1.00 (0.72-1.39) | 1.41 (1.12-1.79) | 1.24 (0.99-1.56) | 1.14 (0.91-1.42) |  | 0.99 (0.90-1.09) | 0.8636 | 0.99 (0.90-1.10) | 0.8812 |
| Model 1, HR (95% CI) | 1.00 (0.71-1.40) | 1.45 (1.14-1.83) | 1.28 (1.03-1.61) | 1.22 (0.97-1.54) |  | 1.01 (0.91-1.12) | 0.8249 | 1.01 (0.91-1.12) | 0.8074 |
| Model 2, HR (95%CI) | 1.00 (0.71-1.41) | 1.47 (1.16-1.86) | 1.31 (1.05-1.64) | 1.27 (1.00-1.61) |  | 1.02 (0.92-1.14) | 0.6736 | 1.02 (0.92-1.14) | 0.6570 |
| Brain (C71) |  |  |  |  |  |  |  |  |  |
| Cases | 39 | 39 | 38 | 47 |  | 163 | | | |
| Model 0, HR (95% CI) | 1.00 (0.72-1.38) | 0.86 (0.63-1.18) | 0.76 (0.55-1.04) | 0.87 (0.65-1.16) |  | 0.97 (0.85-1.09) | 0.5761 | 0.96 (0.85-1.09) | 0.5608 |
| Model 1, HR (95% CI) | 1.00 (0.71-1.40) | 0.85 (0.62-1.16) | 0.71 (0.52-0.98) | 0.77 (0.57-1.06) |  | 0.93 (0.82-1.06) | 0.2995 | 0.93 (0.82-1.06) | 0.2887 |
| Model 2, HR (95%CI) | 1.00 (0.71-1.41) | 0.86 (0.63-1.17) | 0.73 (0.53-0.99) | 0.79 (0.58-1.09) |  | 0.94 (0.82-1.07) | 0.3600 | 0.94 (0.82-1.07) | 0.3479 |
| Non-Hodgkin Lymphoma (C82-85) | |  |  |  |  |  |  |  |  |
| Cases | 67 | 85 | 112 | 105 |  | 369 | | | |
| Model 0, HR (95% CI) | 1.00 (0.78-1.28) | 1.04 (0.84-1.29) | 1.20 (1.00-1.45) | 1.00 (0.82-1.22) |  | 1.00 (0.92-1.08) | 0.9731 | 1.00 (0.92-1.09) | 0.9981 |
| Model 1, HR (95% CI) | 1.00 (0.78-1.29) | 1.05 (0.85-1.29) | 1.22 (1.01-1.46) | 1.03 (0.84-1.26) |  | 1.01 (0.92-1.10) | 0.8762 | 1.01 (0.92-1.10) | 0.8507 |
| Model 2, HR (95%CI) | 1.00 (0.77-1.29) | 1.06 (0.86-1.32) | 1.26 (1.05-1.51) | 1.09 (0.89-1.35) |  | 1.02 (0.94-1.12) | 0.5958 | 1.03 (0.94-1.12) | 0.5735 |
| Diffuse non-Hodgkins lymphoma (C83) | |  |  |  |  |  |  |  |  |
| Cases | 33 | 50 | 56 | 65 |  | 204 | | | |
| Model 0, HR (95% CI) | 1.00 (0.71-1.42) | 1.18 (0.89-1.55) | 1.13 (0.87-1.47) | 1.14 (0.89-1.47) |  | 1.02 (0.92-1.14) | 0.6924 | 1.02 (0.91-1.14) | 0.6872 |
| Model 1, HR (95% CI) | 1.00 (0.70-1.43) | 1.21 (0.92-1.60) | 1.18 (0.91-1.54) | 1.23 (0.95-1.59) |  | 1.04 (0.93-1.17) | 0.4787 | 1.04 (0.93-1.17) | 0.4738 |
| Model 2, HR (95%CI) | 1.00 (0.70-1.43) | 1.23 (0.93-1.62) | 1.22 (0.94-1.57) | 1.30 (0.99-1.69) |  | 1.06 (0.94-1.19) | 0.3465 | 1.06 (0.94-1.20) | 0.3426 |
| Multiple Myeloma (C90) | |  |  |  |  |  |  |  |  |
| Cases | 39 | 29 | 38 | 41 |  | 147 | | | |
| Model 0, HR (95% CI) | 1.00 (0.72-1.39) | 0.59 (0.41-0.84) | 0.67 (0.49-0.92) | 0.63 (0.46-0.86) |  | 0.90 (0.79-1.03) | 0.1312 | 0.90 (0.79-1.03) | 0.1255 |
| Model 1, HR (95% CI) | 1.00 (0.71-1.41) | 0.58 (0.41-0.84) | 0.66 (0.48-0.90) | 0.60 (0.43-0.83) |  | 0.89 (0.78-1.02) | 0.0998 | 0.89 (0.77-1.02) | 0.0953 |
| Model 2, HR (95%CI) | 1.00 (0.71-1.41) | 0.59 (0.41-0.85) | 0.68 (0.50-0.93) | 0.63 (0.45-0.88) |  | 0.91 (0.79-1.04) | 0.1678 | 0.90 (0.78-1.04) | 0.1610 |
| Leukemia (C91-95) |  |  |  |  |  |  |  |  |  |
| Cases | 51 | 69 | 63 | 84 |  | 267 | | | |
| Model 0, HR (95% CI) | 1.00 (0.75-1.33) | 1.06 (0.84-1.34) | 0.85 (0.66-1.09) | 0.98 (0.79-1.23) |  | 0.99 (0.89-1.09) | 0.7671 | 0.98 (0.89-1.09) | 0.7525 |
| Model 1, HR (95% CI) | 1.00 (0.75-1.34) | 1.02 (0.81-1.30) | 0.80 (0.63-1.02) | 0.91 (0.72-1.15) |  | 0.97 (0.87-1.07) | 0.4970 | 0.96 (0.87-1.07) | 0.4826 |
| Model 2, HR (95%CI) | 1.00 (0.75-1.34) | 1.03 (0.81-1.31) | 0.81 (0.63-1.03) | 0.91 (0.72-1.15) |  | 0.96 (0.87-1.07) | 0.4957 | 0.96 (0.87-1.07) | 0.4821 |

Estimates bold if associations are statistically significant (p<0.05).

Model 0: Stratified by age group, geographical region and adjusted for Townsend deprivation score

Model 1: Model 0 + adjusted for racial/ethnic group, height, lives with a spouse or partner, body mass index, cigarette smoking, alcohol consumption, and total physical activity.

Model 2: Model 1 + additionally adjusted for serum concentrations of insulin-like growth factor-I, C-reactive protein, glycated hemoglobin (fourths, unknown).

Abbreviations: CI=Confidence interval; HR=hazard ratio; IQR=interquartile range; SHBG= sex hormone binding globulin.

Supplementary Table S4: HR and 95% CIs for cancer diagnosis by serum free testosterone concentrations in postmenopausal women

|  | Fourths of free testosterone | | | |  | per 10 pmol/L increment | | | |
| --- | --- | --- | --- | --- | --- | --- | --- | --- | --- |
|  |  |  |  |  |  | Not corrected for regression dilution bias | | Corrected for regression dilution bias | |
|  | 1 | 2 | 3 | 4 |  |  | P_trend_ |  | P_trend_ |
| Whole cohort, n | 27657 | 27657 | 27657 | 27657 |  | 110628 | | | |
| Median (IQR), pmol/L | 3.8 (1.9) | 8.3 (2.3) | 13.2 (2.8) | 21.9 (8.6) |  |  |  |  |  |
| Oral (C00-14) |  |  |  |  |  |  |  |  |  |
| Cases | 22 | 19 | 15 | 22 |  | 78 | | | |
| Model 0, HR (95% CI) | 1.00 (0.66-1.52) | 0.88 (0.56-1.37) | 0.67 (0.41-1.12) | 1.01 (0.66-1.53) |  | 1.00 (0.72-1.40) | 0.9913 | 1.01 (0.69-1.49) | 0.9585 |
| Model 1, HR (95% CI) | 1.00 (0.65-1.54) | 0.90 (0.58-1.41) | 0.73 (0.44-1.20) | 1.13 (0.73-1.77) |  | 1.07 (0.75-1.52) | 0.7040 | 1.09 (0.73-1.64) | 0.6738 |
| Model 2, HR (95%CI) | 1.00 (0.65-1.54) | 0.91 (0.58-1.43) | 0.74 (0.44-1.22) | 1.14 (0.73-1.79) |  | 1.08 (0.76-1.53) | 0.6867 | 1.10 (0.73-1.66) | 0.6569 |
| Esophagus (C15) |  |  |  |  |  |  |  |  |  |
| Cases | 20 | 16 | 15 | 12 |  | 63 | | | |
| Model 0, HR (95% CI) | 1.00 (0.64-1.55) | 0.82 (0.50-1.34) | 0.79 (0.48-1.31) | 0.64 (0.36-1.13) |  | 0.79 (0.54-1.16) | 0.2334 | 0.76 (0.48-1.19) | 0.2308 |
| Model 1, HR (95% CI) | 1.00 (0.64-1.57) | 0.86 (0.53-1.41) | 0.85 (0.51-1.40) | 0.68 (0.37-1.22) |  | 0.81 (0.55-1.22) | 0.3182 | 0.79 (0.49-1.26) | 0.3141 |
| Model 2, HR (95%CI) | 1.00 (0.63-1.58) | 0.88 (0.54-1.43) | 0.87 (0.53-1.45) | 0.71 (0.39-1.29) |  | 0.84 (0.56-1.26) | 0.3896 | 0.81 (0.50-1.30) | 0.3841 |
| Adenocarcinoma, esophagus (C15) | |  |  |  |  |  |  |  |  |
| Cases | 9 | 9 | 7 | 9 |  | 34 | | | |
| Model 0, HR (95% CI) | 1.00 (0.52-1.92) | 1.02 (0.53-1.97) | 0.81 (0.39-1.70) | 1.05 (0.55-2.03) |  | 1.01 (0.61-1.67) | 0.9673 | 1.02 (0.57-1.83) | 0.9487 |
| Model 1, HR (95% CI) | 1.00 (0.51-1.95) | 1.02 (0.53-1.97) | 0.74 (0.35-1.55) | 0.84 (0.42-1.68) |  | 0.88 (0.52-1.50) | 0.6419 | 0.87 (0.47-1.61) | 0.6608 |
| Model 2, HR (95%CI) | 1.00 (0.51-1.96) | 1.01 (0.52-1.95) | 0.73 (0.35-1.53) | 0.80 (0.39-1.62) |  | 0.86 (0.51-1.47) | 0.5824 | 0.85 (0.46-1.57) | 0.6004 |
| Stomach (C15) |  |  |  |  |  |  |  |  |  |
| Cases | 9 | 10 | 10 | 13 |  | 42 | | | |
| Model 0, HR (95% CI) | 1.00 (0.52-1.92) | 1.12 (0.60-2.09) | 1.14 (0.61-2.11) | 1.53 (0.89-2.64) |  | 1.26 (0.81-1.96) | 0.3096 | 1.31 (0.78-2.19) | 0.3041 |
| Model 1, HR (95% CI) | 1.00 (0.51-1.96) | 1.13 (0.60-2.11) | 1.21 (0.65-2.24) | 1.68 (0.94-3.00) |  | 1.33 (0.83-2.13) | 0.2315 | 1.40 (0.81-2.41) | 0.2281 |
| Model 2, HR (95%CI) | 1.00 (0.51-1.96) | 1.09 (0.58-2.04) | 1.11 (0.60-2.05) | 1.47 (0.82-2.63) |  | 1.24 (0.77-1.99) | 0.3826 | 1.28 (0.74-2.22) | 0.3770 |
| Colorectum (C18-20) |  |  |  |  |  |  |  |  |  |
| Cases | 163 | 142 | 124 | 138 |  | 567 | | | |
| Model 0, HR (95% CI) | 1.00 (0.86-1.17) | 0.89 (0.76-1.05) | 0.79 (0.66-0.94) | 0.90 (0.76-1.07) |  | 0.94 (0.83-1.07) | 0.3484 | 0.94 (0.81-1.08) | 0.3727 |
| Model 1, HR (95% CI) | 1.00 (0.85-1.17) | 0.88 (0.75-1.04) | 0.76 (0.64-0.91) | 0.85 (0.71-1.01) |  | 0.91 (0.80-1.04) | 0.1492 | 0.90 (0.77-1.04) | 0.1638 |
| Model 2, HR (95%CI) | 1.00 (0.85-1.17) | 0.88 (0.75-1.04) | 0.75 (0.63-0.89) | 0.81 (0.68-0.97) |  | 0.89 (0.78-1.01) | 0.0783 | 0.87 (0.75-1.02) | 0.0877 |
| Colon (C18) |  |  |  |  |  |  |  |  |  |
| Cases | 109 | 104 | 93 | 111 |  | 417 | | | |
| Model 0, HR (95% CI) | 1.00 (0.83-1.21) | 0.98 (0.81-1.19) | 0.89 (0.73-1.09) | 1.09 (0.91-1.32) |  | 1.05 (0.91-1.21) | 0.5343 | 1.06 (0.90-1.25) | 0.5050 |
| Model 1, HR (95% CI) | 1.00 (0.82-1.21) | 0.97 (0.80-1.17) | 0.84 (0.69-1.03) | 0.98 (0.81-1.19) |  | 0.99 (0.85-1.14) | 0.8441 | 0.99 (0.83-1.17) | 0.8831 |
| Model 2, HR (95%CI) | 1.00 (0.82-1.21) | 0.97 (0.80-1.17) | 0.84 (0.68-1.02) | 0.96 (0.79-1.17) |  | 0.97 (0.83-1.13) | 0.7040 | 0.97 (0.81-1.16) | 0.7424 |
| Rectum (C19-20) |  |  |  |  |  |  |  |  |  |
| Cases | 57 | 38 | 31 | 27 |  | 153 | | | |
| Model 0, HR (95% CI) | 1.00 (0.77-1.30) | 0.68 (0.49-0.93) | 0.55 (0.39-0.79) | 0.49 (0.34-0.72) |  | **0.67 (0.52-0.86)** | **0.0020** | **0.62 (0.46-0.84)** | **0.0021** |
| Model 1, HR (95% CI) | 1.00 (0.76-1.31) | 0.67 (0.49-0.92) | 0.55 (0.39-0.79) | 0.50 (0.34-0.74) |  | **0.67 (0.51-0.88)** | **0.0036** | **0.63 (0.46-0.86)** | **0.0037** |
| Model 2, HR (95%CI) | 1.00 (0.76-1.31) | 0.67 (0.49-0.91) | 0.53 (0.38-0.76) | 0.47 (0.31-0.69) |  | **0.64 (0.49-0.84)** | **0.0015** | **0.60 (0.44-0.82)** | **0.0015** |
| Liver (C22) |  |  |  |  |  |  |  |  |  |
| Cases | 15 | 12 | 6 | 15 |  | 48 | | | |
| Model 0, HR (95% CI) | 1.00 (0.60-1.66) | 0.82 (0.47-1.44) | 0.42 (0.19-0.92) | 1.07 (0.65-1.78) |  | 1.03 (0.67-1.57) | 0.8959 | 1.05 (0.64-1.72) | 0.8416 |
| Model 1, HR (95% CI) | 1.00 (0.59-1.69) | 0.83 (0.47-1.45) | 0.43 (0.19-0.96) | 1.06 (0.62-1.83) |  | 1.03 (0.66-1.60) | 0.9067 | 1.05 (0.63-1.75) | 0.8538 |
| Model 2, HR (95%CI) | 1.00 (0.59-1.70) | 0.86 (0.49-1.50) | 0.48 (0.21-1.06) | 1.20 (0.69-2.08) |  | 1.10 (0.70-1.72) | 0.6729 | 1.14 (0.68-1.91) | 0.6272 |
| Pancreas (C25) |  |  |  |  |  |  |  |  |  |
| Cases | 27 | 35 | 27 | 34 |  | 123 | | | |
| Model 0, HR (95% CI) | 1.00 (0.69-1.46) | 1.35 (0.97-1.88) | 1.06 (0.72-1.54) | 1.38 (0.98-1.93) |  | 1.13 (0.87-1.47) | 0.3535 | 1.16 (0.86-1.58) | 0.3262 |
| Model 1, HR (95% CI) | 1.00 (0.68-1.47) | 1.36 (0.98-1.90) | 1.05 (0.72-1.52) | 1.34 (0.94-1.92) |  | 1.11 (0.85-1.46) | 0.4427 | 1.14 (0.83-1.57) | 0.4088 |
| Model 2, HR (95%CI) | 1.00 (0.68-1.47) | 1.37 (0.98-1.91) | 1.05 (0.72-1.53) | 1.36 (0.95-1.95) |  | 1.12 (0.85-1.48) | 0.4240 | 1.15 (0.83-1.59) | 0.3904 |
| Lung (C34) |  |  |  |  |  |  |  |  |  |
| Cases | 96 | 94 | 106 | 111 |  | 407 | | | |
| Model 0, HR (95% CI) | 1.00 (0.82-1.22) | 1.01 (0.83-1.24) | 1.15 (0.95-1.39) | 1.23 (1.02-1.48) |  | 1.13 (0.98-1.31) | 0.0919 | 1.15 (0.98-1.36) | 0.0963 |
| Model 1, HR (95% CI) | 1.00 (0.81-1.23) | 1.03 (0.84-1.27) | 1.17 (0.97-1.41) | 1.23 (1.02-1.50) |  | 1.13 (0.97-1.31) | 0.1092 | 1.15 (0.97-1.37) | 0.1145 |
| Model 2, HR (95%CI) | 1.00 (0.81-1.23) | 1.04 (0.84-1.27) | 1.15 (0.95-1.39) | 1.19 (0.97-1.45) |  | 1.10 (0.95-1.28) | 0.2047 | 1.12 (0.94-1.33) | 0.2125 |
| Melanoma (C43) |  |  |  |  |  |  |  |  |  |
| Cases | 73 | 68 | 59 | 64 |  | 264 | | | |
| Model 0, HR (95% CI) | 1.00 (0.79-1.26) | 0.93 (0.73-1.18) | 0.82 (0.63-1.06) | 0.90 (0.71-1.16) |  | 0.94 (0.78-1.13) | 0.5107 | 0.93 (0.76-1.16) | 0.5312 |
| Model 1, HR (95% CI) | 1.00 (0.79-1.26) | 0.93 (0.73-1.18) | 0.83 (0.64-1.06) | 0.94 (0.73-1.22) |  | 0.96 (0.80-1.17) | 0.7103 | 0.96 (0.77-1.20) | 0.7348 |
| Model 2, HR (95%CI) | 1.00 (0.79-1.27) | 0.94 (0.74-1.19) | 0.84 (0.65-1.09) | 0.97 (0.75-1.26) |  | 0.98 (0.81-1.19) | 0.8485 | 0.98 (0.78-1.23) | 0.8729 |
| Meothelioma (C45) |  |  |  |  |  |  |  |  |  |
| Cases | 7 | 5 | 7 | 4 |  | 23 | | | |
| Model 0, HR (95% CI) | 1.00 (0.48-2.10) | 0.76 (0.32-1.83) | 1.13 (0.54-2.36) | 0.69 (0.26-1.83) |  | 0.88 (0.47-1.65) | 0.6835 | 0.85 (0.40-1.77) | 0.6558 |
| Model 1, HR (95% CI) | 1.00 (0.46-2.17) | 0.85 (0.35-2.04) | 1.38 (0.65-2.95) | 1.04 (0.38-2.86) |  | 1.10 (0.56-2.13) | 0.7846 | 1.10 (0.50-2.39) | 0.8158 |
| Model 2, HR (95%CI) | 1.00 (0.45-2.21) | 0.87 (0.36-2.10) | 1.41 (0.65-3.06) | 1.07 (0.39-2.97) |  | 1.11 (0.56-2.19) | 0.7626 | 1.11 (0.50-2.46) | 0.7932 |
| Breast (C50) |  |  |  |  |  |  |  |  |  |
| Cases | 371 | 400 | 510 | 581 |  | 1862 | | | |
| Model 0, HR (95% CI) | 1.00 (0.90-1.11) | 1.09 (0.99-1.20) | 1.40 (1.29-1.53) | 1.62 (1.50-1.76) |  | **1.32 (1.24-1.41)** | **<0.0001** | **1.38 (1.27-1.49)** | **<0.0001** |
| Model 1, HR (95% CI) | 1.00 (0.90-1.11) | 1.08 (0.98-1.19) | 1.36 (1.25-1.48) | 1.53 (1.40-1.67) |  | **1.28 (1.19-1.37)** | **<0.0001** | **1.32 (1.22-1.43)** | **<0.0001** |
| Model 2, HR (95%CI) | 1.00 (0.90-1.11) | 1.07 (0.97-1.18) | 1.34 (1.23-1.45) | 1.48 (1.36-1.62) |  | **1.25 (1.17-1.34)** | **<0.0001** | **1.29 (1.19-1.40)** | **<0.0001** |
| Endometrial (C54.1) |  |  |  |  |  |  |  |  |  |
| Cases | 66 | 44 | 105 | 155 |  | 370 | | | |
| Model 0, HR (95% CI) | 1.00 (0.79-1.27) | 0.69 (0.51-0.92) | 1.66 (1.37-2.01) | 2.52 (2.15-2.95) |  | **1.88 (1.62-2.18)** | **<0.0001** | **2.05 (1.73-2.44)** | **<0.0001** |
| Model 1, HR (95% CI) | 1.00 (0.78-1.28) | 0.64 (0.48-0.87) | 1.39 (1.15-1.67) | 1.72 (1.45-2.04) |  | **1.50 (1.29-1.76)** | **<0.0001** | **1.59 (1.32-1.90)** | **<0.0001** |
| Model 2, HR (95%CI) | 1.00 (0.78-1.28) | 0.65 (0.48-0.87) | 1.41 (1.17-1.70) | 1.75 (1.48-2.09) |  | **1.52 (1.29-1.78)** | **<0.0001** | **1.60 (1.33-1.92)** | **<0.0001** |
| Ovarian (C56) |  |  |  |  |  |  |  |  |  |
| Cases | 57 | 68 | 50 | 53 |  | 228 | | | |
| Model 0, HR (95% CI) | 1.00 (0.77-1.30) | 1.21 (0.96-1.54) | 0.90 (0.68-1.19) | 0.98 (0.74-1.28) |  | 0.94 (0.77-1.14) | 0.5386 | 0.94 (0.75-1.18) | 0.5845 |
| Model 1, HR (95% CI) | 1.00 (0.77-1.30) | 1.21 (0.95-1.53) | 0.89 (0.68-1.18) | 0.99 (0.74-1.30) |  | 0.95 (0.77-1.16) | 0.5958 | 0.95 (0.75-1.20) | 0.6474 |
| Model 2, HR (95%CI) | 1.00 (0.77-1.31) | 1.22 (0.96-1.55) | 0.92 (0.70-1.21) | 1.04 (0.78-1.38) |  | 0.97 (0.79-1.20) | 0.8098 | 0.98 (0.77-1.25) | 0.8665 |
| Kidney (C64-65) |  |  |  |  |  |  |  |  |  |
| Cases | 21 | 19 | 30 | 32 |  | 102 | | | |
| Model 0, HR (95% CI) | 1.00 (0.65-1.53) | 0.93 (0.59-1.46) | 1.48 (1.03-2.11) | 1.62 (1.14-2.29) |  | **1.36 (1.02-1.81)** | **0.0336** | **1.42 (1.02-1.97)** | **0.0379** |
| Model 1, HR (95% CI) | 1.00 (0.65-1.55) | 0.92 (0.59-1.45) | 1.41 (0.99-2.02) | 1.51 (1.05-2.18) |  | 1.31 (0.97-1.76) | 0.0761 | 1.35 (0.96-1.91) | 0.0844 |
| Model 2, HR (95%CI) | 1.00 (0.64-1.55) | 0.93 (0.59-1.46) | 1.41 (0.99-2.01) | 1.49 (1.03-2.16) |  | 1.29 (0.96-1.75) | 0.0923 | 1.34 (0.94-1.89) | 0.1022 |
| Bladder (C67) |  |  |  |  |  |  |  |  |  |
| Cases | 15 | 15 | 10 | 17 |  | 57 | | | |
| Model 0, HR (95% CI) | 1.00 (0.60-1.66) | 1.03 (0.62-1.70) | 0.69 (0.37-1.29) | 1.22 (0.76-1.96) |  | 1.09 (0.74-1.61) | 0.6485 | 1.12 (0.72-1.76) | 0.6105 |
| Model 1, HR (95% CI) | 1.00 (0.60-1.68) | 1.04 (0.62-1.72) | 0.71 (0.38-1.32) | 1.22 (0.73-2.02) |  | 1.09 (0.73-1.64) | 0.6632 | 1.12 (0.70-1.80) | 0.6257 |
| Model 2, HR (95%CI) | 1.00 (0.59-1.69) | 1.04 (0.63-1.73) | 0.71 (0.38-1.32) | 1.21 (0.72-2.02) |  | 1.09 (0.72-1.64) | 0.6819 | 1.12 (0.70-1.80) | 0.6429 |
| Brain (C71) |  |  |  |  |  |  |  |  |  |
| Cases | 14 | 20 | 21 | 19 |  | 74 | | | |
| Model 0, HR (95% CI) | 1.00 (0.59-1.69) | 1.46 (0.94-2.26) | 1.57 (1.02-2.40) | 1.47 (0.94-2.31) |  | 1.17 (0.84-1.64) | 0.3477 | 1.20 (0.81-1.78) | 0.3514 |
| Model 1, HR (95% CI) | 1.00 (0.59-1.70) | 1.43 (0.92-2.23) | 1.50 (0.98-2.30) | 1.42 (0.89-2.26) |  | 1.15 (0.81-1.64) | 0.4289 | 1.18 (0.78-1.77) | 0.4318 |
| Model 2, HR (95%CI) | 1.00 (0.59-1.71) | 1.45 (0.93-2.25) | 1.53 (1.00-2.35) | 1.46 (0.91-2.34) |  | 1.17 (0.82-1.67) | 0.3883 | 1.20 (0.79-1.82) | 0.3913 |
| Non-Hodgkin Lymphoma (C82-85) | |  |  |  |  |  |  |  |  |
| Cases | 62 | 63 | 45 | 39 |  | 209 | | | |
| Model 0, HR (95% CI) | 1.00 (0.78-1.28) | 1.04 (0.81-1.32) | 0.76 (0.56-1.01) | 0.67 (0.49-0.92) |  | **0.78 (0.63-0.96)** | **0.0201** | **0.75 (0.58-0.96)** | **0.0230** |
| Model 1, HR (95% CI) | 1.00 (0.77-1.29) | 1.03 (0.80-1.32) | 0.76 (0.56-1.01) | 0.69 (0.50-0.95) |  | **0.79 (0.63-0.98)** | **0.0353** | **0.76 (0.59-0.99)** | **0.0401** |
| Model 2, HR (95%CI) | 1.00 (0.77-1.29) | 1.05 (0.82-1.34) | 0.78 (0.58-1.04) | 0.71 (0.51-0.98) |  | 0.80 (0.64-1.00) | 0.0517 | 0.78 (0.60-1.01) | 0.0584 |
| Diffuse non-Hodgkins lymphoma (C83) | |  |  |  |  |  |  |  |  |
| Cases | 29 | 33 | 20 | 24 |  | 106 | | | |
| Model 0, HR (95% CI) | 1.00 (0.69-1.44) | 1.16 (0.82-1.63) | 0.71 (0.46-1.11) | 0.88 (0.59-1.32) |  | 0.88 (0.66-1.18) | 0.3875 | 0.87 (0.62-1.23) | 0.4287 |
| Model 1, HR (95% CI) | 1.00 (0.69-1.45) | 1.14 (0.81-1.61) | 0.69 (0.45-1.07) | 0.87 (0.57-1.32) |  | 0.87 (0.64-1.18) | 0.3690 | 0.86 (0.60-1.23) | 0.4120 |
| Model 2, HR (95%CI) | 1.00 (0.69-1.45) | 1.16 (0.82-1.63) | 0.69 (0.45-1.08) | 0.84 (0.55-1.28) |  | 0.85 (0.63-1.16) | 0.3153 | 0.84 (0.59-1.21) | 0.3547 |
| Multiple Myeloma (C90) | |  |  |  |  |  |  |  |  |
| Cases | 28 | 19 | 17 | 18 |  | 82 | | | |
| Model 0, HR (95% CI) | 1.00 (0.69-1.45) | 0.71 (0.45-1.11) | 0.64 (0.40-1.03) | 0.69 (0.43-1.10) |  | 0.82 (0.58-1.14) | 0.2406 | 0.79 (0.54-1.17) | 0.2437 |
| Model 1, HR (95% CI) | 1.00 (0.68-1.47) | 0.67 (0.43-1.04) | 0.55 (0.34-0.89) | 0.50 (0.31-0.82) |  | **0.69 (0.48-0.97)** | **0.0339** | **0.65 (0.43-0.97)** | **0.0348** |
| Model 2, HR (95%CI) | 1.00 (0.68-1.47) | 0.67 (0.43-1.05) | 0.56 (0.35-0.90) | 0.51 (0.31-0.83) |  | **0.69 (0.48-0.98)** | **0.0385** | **0.65 (0.43-0.98)** | **0.0396** |
| Leukemia (C91-95) |  |  |  |  |  |  |  |  |  |
| Cases | 32 | 31 | 24 | 26 |  | 113 | | | |
| Model 0, HR (95% CI) | 1.00 (0.71-1.42) | 0.99 (0.70-1.41) | 0.80 (0.53-1.19) | 0.89 (0.61-1.31) |  | 0.92 (0.69-1.22) | 0.5596 | 0.91 (0.66-1.27) | 0.5824 |
| Model 1, HR (95% CI) | 1.00 (0.70-1.43) | 1.02 (0.72-1.46) | 0.84 (0.56-1.25) | 0.93 (0.63-1.40) |  | 0.94 (0.70-1.26) | 0.6956 | 0.94 (0.67-1.32) | 0.7196 |
| Model 2, HR (95%CI) | 1.00 (0.70-1.43) | 1.03 (0.73-1.47) | 0.84 (0.56-1.25) | 0.94 (0.63-1.41) |  | 0.95 (0.70-1.27) | 0.7078 | 0.94 (0.67-1.33) | 0.7335 |

Estimates bold if associations are statistically significant (p<0.05).

Model 0: Stratified by age group, geographical region and adjusted for Townsend deprivation score

Model 1: Model 0 + adjusted for racial/ethnic group, height, lives with a spouse or partner, body mass index, cigarette smoking, alcohol consumption, total physical activity, hormone replacement therapy use, oral contraceptive use, and parity and age at first birth

Model 2: Model 1 + additionally adjusted for serum concentrations of insulin-like growth factor-I, C-reactive protein, glycated hemoglobin (fourths, unknown).

Abbreviations: CI=Confidence interval; HR=hazard ratio; IQR=interquartile range.

Supplementary Table S5: HR and 95% CIs for cancer diagnosis by serum total testosterone concentrations in postmenopausal women

|  | Fourths of total testosterone | | | |  | per 0.5 nmol/L increment | | | |
| --- | --- | --- | --- | --- | --- | --- | --- | --- | --- |
|  |  |  |  |  |  | Not corrected for regression dilution bias | | Corrected for regression dilution bias | |
|  | 1 | 2 | 3 | 4 |  |  | P_trend_ |  | P_trend_ |
| Whole cohort, n | 30557 | 30542 | 30514 | 30499 |  | 122112 | | | |
| Median (IQR), nmol/L | 0.3 (0.1) | 0.7 (0.2) | 1.0 (0.2) | 1.6 (0.5) |  |  |  |  |  |
| Oral (C00-14) |  |  |  |  |  |  |  |  |  |
| Cases | 24 | 16 | 21 | 20 |  | 81 | | | |
| Model 0, HR (95% CI) | 1.00 (0.67-1.49) | 0.67 (0.41-1.09) | 0.88 (0.57-1.35) | 0.84 (0.54-1.31) |  | 0.96 (0.76-1.21) | 0.7413 | 0.96 (0.72-1.27) | 0.7586 |
| Model 1, HR (95% CI) | 1.00 (0.67-1.50) | 0.69 (0.43-1.13) | 0.91 (0.59-1.40) | 0.86 (0.55-1.35) |  | 0.97 (0.77-1.22) | 0.7940 | 0.97 (0.73-1.28) | 0.8073 |
| Model 2, HR (95%CI) | 1.00 (0.67-1.50) | 0.70 (0.43-1.14) | 0.91 (0.59-1.40) | 0.86 (0.55-1.35) |  | 0.97 (0.77-1.22) | 0.7811 | 0.96 (0.72-1.28) | 0.7938 |
| Esophagus (C15) |  |  |  |  |  |  |  |  |  |
| Cases | 22 | 20 | 11 | 16 |  | 69 | | | |
| Model 0, HR (95% CI) | 1.00 (0.66-1.52) | 0.94 (0.61-1.47) | 0.53 (0.29-0.96) | 0.79 (0.48-1.28) |  | 0.87 (0.67-1.12) | 0.2720 | 0.85 (0.62-1.16) | 0.2974 |
| Model 1, HR (95% CI) | 1.00 (0.65-1.53) | 0.98 (0.63-1.52) | 0.54 (0.30-0.98) | 0.79 (0.48-1.30) |  | 0.87 (0.67-1.12) | 0.2846 | 0.85 (0.62-1.16) | 0.3091 |
| Model 2, HR (95%CI) | 1.00 (0.65-1.53) | 0.97 (0.63-1.51) | 0.54 (0.30-0.98) | 0.80 (0.48-1.31) |  | 0.87 (0.68-1.13) | 0.2989 | 0.85 (0.62-1.17) | 0.3244 |
| Adenocarcinoma, esophagus (C15) | |  |  |  |  |  |  |  |  |
| Cases | 13 | 7 | 6 | 8 |  | 34 | | | |
| Model 0, HR (95% CI) | 1.00 (0.58-1.72) | 0.56 (0.27-1.18) | 0.49 (0.22-1.08) | 0.66 (0.33-1.33) |  | 0.83 (0.58-1.19) | 0.3091 | 0.80 (0.51-1.26) | 0.3391 |
| Model 1, HR (95% CI) | 1.00 (0.57-1.75) | 0.56 (0.27-1.17) | 0.46 (0.21-1.02) | 0.60 (0.30-1.21) |  | 0.79 (0.55-1.15) | 0.2213 | 0.76 (0.49-1.20) | 0.2455 |
| Model 2, HR (95%CI) | 1.00 (0.57-1.75) | 0.56 (0.27-1.18) | 0.46 (0.20-1.02) | 0.59 (0.29-1.19) |  | 0.79 (0.55-1.14) | 0.2075 | 0.76 (0.48-1.19) | 0.2302 |
| Stomach (C15) |  |  |  |  |  |  |  |  |  |
| Cases | 6 | 11 | 16 | 13 |  | 46 | | | |
| Model 0, HR (95% CI) | 1.00 (0.45-2.23) | 1.88 (1.04-3.39) | 2.81 (1.72-4.59) | 2.32 (1.35-4.00) |  | 1.32 (0.97-1.80) | 0.0730 | 1.38 (0.96-2.00) | 0.0842 |
| Model 1, HR (95% CI) | 1.00 (0.45-2.24) | 1.92 (1.06-3.47) | 2.88 (1.76-4.71) | 2.46 (1.42-4.27) |  | 1.35 (0.99-1.85) | 0.0577 | 1.42 (0.98-2.06) | 0.0669 |
| Model 2, HR (95%CI) | 1.00 (0.45-2.24) | 1.90 (1.05-3.43) | 2.77 (1.69-4.53) | 2.28 (1.31-3.97) |  | 1.31 (0.96-1.79) | 0.0897 | 1.37 (0.94-1.99) | 0.1034 |
| Colorectum (C18-20) |  |  |  |  |  |  |  |  |  |
| Cases | 172 | 144 | 149 | 145 |  | 129 | | | |
| Model 0, HR (95% CI) | 1.00 (0.86-1.16) | 0.86 (0.73-1.02) | 0.91 (0.77-1.07) | 0.89 (0.76-1.05) |  | 0.96 (0.89-1.05) | 0.3830 | 0.96 (0.86-1.06) | 0.3988 |
| Model 1, HR (95% CI) | 1.00 (0.86-1.16) | 0.86 (0.73-1.01) | 0.90 (0.76-1.05) | 0.87 (0.74-1.02) |  | 0.95 (0.88-1.04) | 0.2753 | 0.95 (0.85-1.05) | 0.2880 |
| Model 2, HR (95%CI) | 1.00 (0.86-1.16) | 0.86 (0.73-1.01) | 0.89 (0.76-1.05) | 0.86 (0.73-1.02) |  | 0.95 (0.87-1.04) | 0.2457 | 0.94 (0.85-1.05) | 0.2572 |
| Colon (C18) |  |  |  |  |  |  |  |  |  |
| Cases | 121 | 104 | 103 | 117 |  | 445 | | | |
| Model 0, HR (95% CI) | 1.00 (0.84-1.20) | 0.89 (0.74-1.08) | 0.90 (0.74-1.09) | 1.03 (0.86-1.24) |  | 1.01 (0.92-1.12) | 0.7712 | 1.02 (0.91-1.15) | 0.7270 |
| Model 1, HR (95% CI) | 1.00 (0.83-1.20) | 0.88 (0.73-1.07) | 0.88 (0.73-1.07) | 0.99 (0.82-1.19) |  | 1.00 (0.90-1.10) | 0.9590 | 1.00 (0.89-1.13) | 0.9948 |
| Model 2, HR (95%CI) | 1.00 (0.83-1.20) | 0.88 (0.73-1.07) | 0.88 (0.72-1.06) | 0.98 (0.81-1.18) |  | 0.99 (0.90-1.10) | 0.9057 | 1.00 (0.88-1.12) | 0.9512 |
| Rectum (C19-20) |  |  |  |  |  |  |  |  |  |
| Cases | 53 | 41 | 46 | 28 |  | 168 | | | |
| Model 0, HR (95% CI) | 1.00 (0.76-1.31) | 0.78 (0.58-1.07) | 0.89 (0.66-1.19) | 0.54 (0.38-0.79) |  | **0.82 (0.70-0.97)** | **0.0183** | **0.78 (0.64-0.96)** | **0.0167** |
| Model 1, HR (95% CI) | 1.00 (0.76-1.31) | 0.79 (0.58-1.07) | 0.89 (0.66-1.18) | 0.55 (0.38-0.79) |  | **0.82 (0.70-0.97)** | **0.0206** | **0.78 (0.64-0.96)** | **0.0188** |
| Model 2, HR (95%CI) | 1.00 (0.76-1.32) | 0.79 (0.58-1.07) | 0.89 (0.67-1.19) | 0.55 (0.38-0.79) |  | **0.82 (0.70-0.97)** | **0.0206** | **0.78 (0.64-0.96)** | **0.0187** |
| Liver (C22) |  |  |  |  |  |  |  |  |  |
| Cases | 15 | 10 | 8 | 21 |  | 54 | | | |
| Model 0, HR (95% CI) | 1.00 (0.60-1.66) | 0.69 (0.37-1.27) | 0.56 (0.28-1.12) | 1.51 (0.98-2.32) |  | 1.21 (0.91-1.60) | 0.1884 | 1.28 (0.91-1.80) | 0.1520 |
| Model 1, HR (95% CI) | 1.00 (0.60-1.67) | 0.72 (0.39-1.33) | 0.58 (0.29-1.15) | 1.55 (1.00-2.40) |  | 1.22 (0.92-1.61) | 0.1772 | 1.29 (0.92-1.82) | 0.1433 |
| Model 2, HR (95%CI) | 1.00 (0.60-1.68) | 0.73 (0.39-1.36) | 0.60 (0.30-1.20) | 1.69 (1.09-2.62) |  | 1.26 (0.94-1.67) | 0.1190 | 1.34 (0.95-1.90) | 0.0935 |
| Pancreas (C25) |  |  |  |  |  |  |  |  |  |
| Cases | 25 | 40 | 31 | 43 |  | 139 | | | |
| Model 0, HR (95% CI) | 1.00 (0.68-1.48) | 1.68 (1.23-2.30) | 1.33 (0.94-1.90) | 1.89 (1.40-2.55) |  | **1.22 (1.02-1.45)** | **0.0282** | **1.27 (1.03-1.57)** | **0.0277** |
| Model 1, HR (95% CI) | 1.00 (0.67-1.49) | 1.69 (1.24-2.30) | 1.33 (0.94-1.89) | 1.85 (1.37-2.51) |  | **1.21 (1.01-1.44)** | **0.0387** | **1.25 (1.01-1.55)** | **0.0385** |
| Model 2, HR (95%CI) | 1.00 (0.67-1.49) | 1.69 (1.24-2.30) | 1.34 (0.94-1.90) | 1.85 (1.37-2.51) |  | **1.21 (1.01-1.44)** | **0.0392** | **1.25 (1.01-1.55)** | **0.0391** |
| Lung (C34) |  |  |  |  |  |  |  |  |  |
| Cases | 111 | 115 | 114 | 113 |  | 453 | | | |
| Model 0, HR (95% CI) | 1.00 (0.83-1.20) | 1.09 (0.91-1.31) | 1.10 (0.92-1.33) | 1.11 (0.93-1.34) |  | 1.04 (0.94-1.14) | 0.4350 | 1.05 (0.93-1.18) | 0.4464 |
| Model 1, HR (95% CI) | 1.00 (0.83-1.21) | 1.11 (0.92-1.33) | 1.07 (0.89-1.29) | 1.03 (0.85-1.24) |  | 1.01 (0.91-1.11) | 0.9140 | 1.00 (0.89-1.13) | 0.9387 |
| Model 2, HR (95%CI) | 1.00 (0.83-1.21) | 1.12 (0.93-1.34) | 1.08 (0.90-1.30) | 1.03 (0.86-1.24) |  | 1.01 (0.91-1.11) | 0.8925 | 1.01 (0.89-1.13) | 0.9199 |
| Melanoma (C43) |  |  |  |  |  |  |  |  |  |
| Cases | 88 | 60 | 73 | 71 |  | 292 | | | |
| Model 0, HR (95% CI) | 1.00 (0.81-1.23) | 0.69 (0.54-0.89) | 0.85 (0.68-1.07) | 0.84 (0.66-1.06) |  | 0.95 (0.84-1.08) | 0.4364 | 0.95 (0.82-1.10) | 0.4654 |
| Model 1, HR (95% CI) | 1.00 (0.81-1.24) | 0.68 (0.53-0.88) | 0.85 (0.67-1.07) | 0.84 (0.67-1.06) |  | 0.95 (0.84-1.08) | 0.4580 | 0.95 (0.82-1.10) | 0.4899 |
| Model 2, HR (95%CI) | 1.00 (0.81-1.24) | 0.69 (0.53-0.89) | 0.85 (0.68-1.07) | 0.85 (0.67-1.08) |  | 0.96 (0.85-1.09) | 0.5079 | 0.95 (0.82-1.11) | 0.5417 |
| Meothelioma (C45) |  |  |  |  |  |  |  |  |  |
| Cases | 4 | 9 | 11 | 2 |  | 26 | | | |
| Model 0, HR (95% CI) | 1.00 (0.38-2.67) | 2.41 (1.25-4.63) | 3.08 (1.71-5.57) | 0.58 (0.14-2.32) |  | 0.95 (0.63-1.43) | 0.8189 | 0.91 (0.55-1.50) | 0.7147 |
| Model 1, HR (95% CI) | 1.00 (0.37-2.68) | 2.75 (1.42-5.33) | 3.34 (1.81-6.16) | 0.72 (0.18-2.91) |  | 1.03 (0.68-1.56) | 0.9039 | 1.00 (0.60-1.67) | 0.9879 |
| Model 2, HR (95%CI) | 1.00 (0.37-2.70) | 2.65 (1.36-5.18) | 3.34 (1.81-6.17) | 0.73 (0.18-2.94) |  | 1.03 (0.67-1.57) | 0.8940 | 1.00 (0.59-1.68) | 0.9982 |
| Breast (C50) |  |  |  |  |  |  |  |  |  |
| Cases | 409 | 472 | 523 | 637 |  | 2041 | | | |
| Model 0, HR (95% CI) | 1.00 (0.91-1.10) | 1.17 (1.07-1.28) | 1.32 (1.21-1.44) | 1.63 (1.51-1.76) |  | **1.21 (1.15-1.26)** | **<0.0001** | **1.26 (1.19-1.33)** | **<0.0001** |
| Model 1, HR (95% CI) | 1.00 (0.91-1.10) | 1.16 (1.06-1.27) | 1.29 (1.19-1.41) | 1.58 (1.46-1.71) |  | **1.19 (1.14-1.25)** | **<0.0001** | **1.24 (1.17-1.31)** | **<0.0001** |
| Model 2, HR (95%CI) | 1.00 (0.91-1.10) | 1.16 (1.06-1.27) | 1.28 (1.18-1.39) | 1.55 (1.43-1.68) |  | **1.18 (1.13-1.24)** | **<0.0001** | **1.23 (1.16-1.30)** | **<0.0001** |
| Endometrial (C54.1) |  |  |  |  |  |  |  |  |  |
| Cases | 67 | 85 | 105 | 145 |  | 402 | | | |
| Model 0, HR (95% CI) | 1.00 (0.79-1.27) | 1.31 (1.06-1.63) | 1.65 (1.36-2.00) | 2.33 (1.98-2.75) |  | **1.39 (1.25-1.54)** | **<0.0001** | **1.49 (1.31-1.69)** | **<0.0001** |
| Model 1, HR (95% CI) | 1.00 (0.79-1.27) | 1.23 (1.00-1.53) | 1.47 (1.22-1.78) | 1.87 (1.58-2.20) |  | **1.27 (1.15-1.41)** | **<0.0001** | **1.34 (1.18-1.52)** | **<0.0001** |
| Model 2, HR (95%CI) | 1.00 (0.79-1.27) | 1.24 (1.00-1.54) | 1.49 (1.23-1.80) | 1.90 (1.61-2.25) |  | **1.28 (1.15-1.42)** | **<0.0001** | **1.35 (1.19-1.53)** | **<0.0001** |
| Ovarian (C56) |  |  |  |  |  |  |  |  |  |
| Cases | 59 | 83 | 54 | 57 |  | 253 | | | |
| Model 0, HR (95% CI) | 1.00 (0.77-1.29) | 1.44 (1.16-1.79) | 0.95 (0.73-1.24) | 1.02 (0.78-1.32) |  | 0.96 (0.84-1.10) | 0.5562 | 0.95 (0.81-1.12) | 0.5419 |
| Model 1, HR (95% CI) | 1.00 (0.77-1.29) | 1.45 (1.17-1.79) | 0.95 (0.72-1.23) | 1.03 (0.79-1.33) |  | 0.96 (0.84-1.10) | 0.5901 | 0.95 (0.81-1.12) | 0.5767 |
| Model 2, HR (95%CI) | 1.00 (0.77-1.29) | 1.45 (1.17-1.80) | 0.95 (0.73-1.25) | 1.05 (0.81-1.37) |  | 0.97 (0.85-1.11) | 0.6864 | 0.97 (0.82-1.14) | 0.6728 |
| Kidney (C64-65) |  |  |  |  |  |  |  |  |  |
| Cases | 24 | 34 | 22 | 35 |  | 115 | | | |
| Model 0, HR (95% CI) | 1.00 (0.67-1.49) | 1.46 (1.04-2.04) | 0.96 (0.63-1.46) | 1.55 (1.11-2.16) |  | 1.13 (0.93-1.37) | 0.2092 | 1.17 (0.92-1.47) | 0.1961 |
| Model 1, HR (95% CI) | 1.00 (0.67-1.50) | 1.44 (1.03-2.02) | 0.93 (0.61-1.41) | 1.44 (1.03-2.02) |  | 1.10 (0.90-1.33) | 0.3504 | 1.12 (0.89-1.42) | 0.3328 |
| Model 2, HR (95%CI) | 1.00 (0.67-1.50) | 1.46 (1.04-2.04) | 0.94 (0.62-1.42) | 1.44 (1.03-2.02) |  | 1.10 (0.90-1.33) | 0.3520 | 1.12 (0.89-1.42) | 0.3357 |
| Bladder (C67) |  |  |  |  |  |  |  |  |  |
| Cases | 14 | 14 | 17 | 15 |  | 60 | | | |
| Model 0, HR (95% CI) | 1.00 (0.59-1.69) | 1.04 (0.62-1.76) | 1.29 (0.80-2.08) | 1.15 (0.69-1.90) |  | 1.07 (0.82-1.40) | 0.6192 | 1.08 (0.78-1.49) | 0.6359 |
| Model 1, HR (95% CI) | 1.00 (0.59-1.70) | 1.06 (0.63-1.79) | 1.29 (0.80-2.08) | 1.11 (0.66-1.85) |  | 1.05 (0.81-1.38) | 0.7044 | 1.06 (0.76-1.47) | 0.7253 |
| Model 2, HR (95%CI) | 1.00 (0.59-1.70) | 1.06 (0.63-1.79) | 1.28 (0.79-2.05) | 1.11 (0.66-1.85) |  | 1.05 (0.80-1.38) | 0.7080 | 1.06 (0.76-1.47) | 0.7275 |
| Brain (C71) |  |  |  |  |  |  |  |  |  |
| Cases | 17 | 24 | 21 | 20 |  | 82 | | | |
| Model 0, HR (95% CI) | 1.00 (0.62-1.61) | 1.45 (0.97-2.16) | 1.31 (0.85-2.00) | 1.29 (0.83-1.99) |  | 1.07 (0.85-1.35) | 0.5496 | 1.08 (0.82-1.43) | 0.5710 |
| Model 1, HR (95% CI) | 1.00 (0.62-1.61) | 1.40 (0.94-2.09) | 1.25 (0.82-1.92) | 1.22 (0.79-1.91) |  | 1.05 (0.84-1.33) | 0.6631 | 1.06 (0.80-1.41) | 0.6845 |
| Model 2, HR (95%CI) | 1.00 (0.62-1.62) | 1.40 (0.94-2.09) | 1.25 (0.81-1.91) | 1.21 (0.78-1.89) |  | 1.05 (0.83-1.32) | 0.6887 | 1.05 (0.80-1.40) | 0.7110 |
| Non-Hodgkin Lymphoma (C82-85) | |  |  |  |  |  |  |  |  |
| Cases | 70 | 60 | 45 | 60 |  | 235 | | | |
| Model 0, HR (95% CI) | 1.00 (0.79-1.26) | 0.88 (0.68-1.13) | 0.67 (0.50-0.90) | 0.91 (0.70-1.17) |  | 0.94 (0.82-1.08) | 0.3872 | 0.94 (0.79-1.11) | 0.4382 |
| Model 1, HR (95% CI) | 1.00 (0.79-1.27) | 0.88 (0.68-1.13) | 0.68 (0.51-0.91) | 0.93 (0.72-1.20) |  | 0.95 (0.83-1.09) | 0.4702 | 0.95 (0.80-1.12) | 0.5283 |
| Model 2, HR (95%CI) | 1.00 (0.79-1.27) | 0.89 (0.69-1.15) | 0.69 (0.51-0.92) | 0.94 (0.73-1.22) |  | 0.96 (0.83-1.10) | 0.5399 | 0.96 (0.81-1.13) | 0.6011 |
| Diffuse non-Hodgkins lymphoma (C83) | |  |  |  |  |  |  |  |  |
| Cases | 31 | 27 | 22 | 35 |  | 115 | | | |
| Model 0, HR (95% CI) | 1.00 (0.70-1.42) | 0.89 (0.61-1.30) | 0.75 (0.49-1.13) | 1.19 (0.85-1.66) |  | 1.06 (0.88-1.29) | 0.5343 | 1.09 (0.86-1.37) | 0.4837 |
| Model 1, HR (95% CI) | 1.00 (0.70-1.43) | 0.88 (0.61-1.29) | 0.75 (0.49-1.14) | 1.20 (0.86-1.68) |  | 1.07 (0.88-1.30) | 0.5060 | 1.09 (0.86-1.39) | 0.4554 |
| Model 2, HR (95%CI) | 1.00 (0.70-1.43) | 0.90 (0.62-1.31) | 0.75 (0.50-1.15) | 1.22 (0.87-1.70) |  | 1.07 (0.88-1.30) | 0.4875 | 1.10 (0.87-1.39) | 0.4386 |
| Multiple Myeloma (C90) | |  |  |  |  |  |  |  |  |
| Cases | 35 | 18 | 25 | 18 |  | 96 | | | |
| Model 0, HR (95% CI) | 1.00 (0.72-1.39) | 0.54 (0.34-0.86) | 0.77 (0.52-1.14) | 0.55 (0.34-0.87) |  | 0.82 (0.66-1.02) | 0.0702 | 0.78 (0.60-1.02) | 0.0731 |
| Model 1, HR (95% CI) | 1.00 (0.71-1.40) | 0.51 (0.32-0.82) | 0.70 (0.47-1.03) | 0.47 (0.30-0.76) |  | **0.77 (0.62-0.96)** | **0.0214** | **0.73 (0.56-0.96)** | **0.0228** |
| Model 2, HR (95%CI) | 1.00 (0.71-1.40) | 0.51 (0.32-0.81) | 0.69 (0.47-1.02) | 0.47 (0.29-0.75) |  | **0.77 (0.62-0.96)** | **0.0193** | **0.73 (0.55-0.95)** | **0.0206** |
| Leukemia (C91-95) |  |  |  |  |  |  |  |  |  |
| Cases | 32 | 30 | 32 | 35 |  | 129 | | | |
| Model 0, HR (95% CI) | 1.00 (0.71-1.41) | 0.98 (0.68-1.40) | 1.07 (0.75-1.51) | 1.19 (0.86-1.66) |  | 1.08 (0.90-1.29) | 0.4207 | 1.10 (0.88-1.37) | 0.4128 |
| Model 1, HR (95% CI) | 1.00 (0.70-1.42) | 1.00 (0.70-1.44) | 1.11 (0.79-1.57) | 1.27 (0.91-1.77) |  | 1.10 (0.92-1.32) | 0.3005 | 1.13 (0.90-1.41) | 0.2944 |
| Model 2, HR (95%CI) | 1.00 (0.70-1.42) | 1.01 (0.70-1.44) | 1.10 (0.78-1.56) | 1.25 (0.90-1.76) |  | 1.10 (0.91-1.32) | 0.3253 | 1.12 (0.90-1.40) | 0.3192 |

Estimates bold if associations are statistically significant (p<0.05).

Model 0: Stratified by age group, geographical region and adjusted for Townsend deprivation score

Model 1: Model 0 + adjusted for racial/ethnic group, height, lives with a spouse or partner, body mass index, cigarette smoking, alcohol consumption, total physical activity, hormone replacement therapy use, oral contraceptive use, and parity and age at first birth

Model 2: Model 1 + additionally adjusted for serum concentrations of insulin-like growth factor-I, C-reactive protein, glycated hemoglobin (fourths, unknown).

Abbreviations: CI=Confidence interval; HR=hazard ratio; IQR=interquartile range.

Supplementary Table S6: HR and 95% CIs for cancer diagnosis by serum SHBG concentrations in postmenopausal women

|  | Fourths of SHBG | | | |  | per 25 nmol/L increment | | | |
| --- | --- | --- | --- | --- | --- | --- | --- | --- | --- |
|  |  |  |  |  |  | Not corrected for regression dilution bias | | Corrected for regression dilution bias | |
|  | 1 | 2 | 3 | 4 |  |  | P_trend_ |  | P_trend_ |
| Whole cohort, n | 27809 | 27780 | 27794 | 27787 |  | 111170 | | |  |
| Median (IQR), nmol/L | 31.3 (10.0) | 47.2 (7.4) | 63.0 (9.2) | 88.9 (23.3) |  |  |  |  |  |
| Oral (C00-14) |  |  |  |  |  |  |  |  |  |
| Cases | 22 | 18 | 14 | 24 |  | 78 | | | |
| Model 0, HR (95% CI) | 1.00 (0.66-1.52) | 0.83 (0.52-1.32) | 0.66 (0.39-1.11) | 1.13 (0.75-1.68) |  | 1.06 (0.82-1.37) | 0.6678 | 1.06 (0.81-1.40) | 0.6617 |
| Model 1, HR (95% CI) | 1.00 (0.63-1.59) | 0.78 (0.49-1.23) | 0.58 (0.35-0.98) | 0.91 (0.59-1.43) |  | 0.98 (0.73-1.30) | 0.8713 | 0.98 (0.72-1.33) | 0.8785 |
| Model 2, HR (95%CI) | 1.00 (0.62-1.62) | 0.77 (0.49-1.22) | 0.58 (0.34-0.97) | 0.89 (0.56-1.42) |  | 0.97 (0.71-1.31) | 0.8264 | 0.97 (0.70-1.33) | 0.8337 |
| Esophagus (C15) |  |  |  |  |  |  |  |  |  |
| Cases | 15 | 16 | 13 | 20 |  | 64 | | | |
| Model 0, HR (95% CI) | 1.00 (0.60-1.66) | 1.07 (0.65-1.74) | 0.86 (0.50-1.48) | 1.32 (0.85-2.04) |  | 1.12 (0.84-1.48) | 0.4535 | 1.12 (0.83-1.52) | 0.4493 |
| Model 1, HR (95% CI) | 1.00 (0.57-1.75) | 1.05 (0.65-1.71) | 0.82 (0.48-1.42) | 1.23 (0.76-2.00) |  | 1.09 (0.79-1.50) | 0.6021 | 1.09 (0.78-1.53) | 0.5967 |
| Model 2, HR (95%CI) | 1.00 (0.56-1.77) | 1.03 (0.63-1.68) | 0.80 (0.47-1.37) | 1.15 (0.69-1.91) |  | 1.06 (0.76-1.47) | 0.7516 | 1.06 (0.75-1.51) | 0.7455 |
| Adenocarcinoma, esophagus (C15) | |  |  |  |  |  |  |  |  |
| Cases | 14 | 6 | 8 | 7 |  | 35 | | | |
| Model 0, HR (95% CI) | 1.00 (0.59-1.69) | 0.44 (0.20-0.97) | 0.58 (0.29-1.15) | 0.51 (0.24-1.07) |  | 0.76 (0.51-1.15) | 0.1963 | 0.75 (0.49-1.16) | 0.1952 |
| Model 1, HR (95% CI) | 1.00 (0.54-1.84) | 0.49 (0.22-1.08) | 0.73 (0.37-1.47) | 0.69 (0.31-1.54) |  | 0.88 (0.56-1.39) | 0.5938 | 0.88 (0.54-1.42) | 0.5903 |
| Model 2, HR (95%CI) | 1.00 (0.53-1.90) | 0.50 (0.23-1.09) | 0.75 (0.38-1.50) | 0.72 (0.32-1.65) |  | 0.90 (0.56-1.45) | 0.6679 | 0.89 (0.54-1.48) | 0.6639 |
| Stomach (C15) |  |  |  |  |  |  |  |  |  |
| Cases | 13 | 11 | 6 | 12 |  | 42 | | | |
| Model 0, HR (95% CI) | 1.00 (0.58-1.73) | 0.83 (0.46-1.51) | 0.46 (0.21-1.02) | 0.93 (0.53-1.64) |  | 0.94 (0.66-1.35) | 0.7440 | 0.94 (0.64-1.38) | 0.7517 |
| Model 1, HR (95% CI) | 1.00 (0.54-1.84) | 0.82 (0.46-1.46) | 0.43 (0.19-0.95) | 0.80 (0.43-1.51) |  | 0.89 (0.60-1.33) | 0.5748 | 0.89 (0.58-1.36) | 0.5825 |
| Model 2, HR (95%CI) | 1.00 (0.53-1.87) | 0.84 (0.47-1.50) | 0.47 (0.21-1.04) | 0.95 (0.49-1.84) |  | 0.96 (0.64-1.47) | 0.8661 | 0.97 (0.62-1.50) | 0.8753 |
| Colorectum (C18-20) |  |  |  |  |  |  |  |  |  |
| Cases | 150 | 123 | 141 | 158 |  | 572 | | | |
| Model 0, HR (95% CI) | 1.00 (0.85-1.17) | 0.79 (0.66-0.95) | 0.90 (0.77-1.07) | 1.01 (0.86-1.18) |  | 1.03 (0.94-1.14) | 0.5306 | 1.03 (0.93-1.14) | 0.5322 |
| Model 1, HR (95% CI) | 1.00 (0.84-1.19) | 0.83 (0.69-0.98) | 0.96 (0.82-1.13) | 1.11 (0.94-1.31) |  | 1.07 (0.97-1.20) | 0.1857 | 1.08 (0.96-1.21) | 0.1867 |
| Model 2, HR (95%CI) | 1.00 (0.83-1.20) | 0.86 (0.72-1.02) | 1.03 (0.88-1.22) | 1.24 (1.04-1.49) |  | **1.13 (1.01-1.26)** | **0.0328** | **1.14 (1.01-1.28)** | **0.0331** |
| Colon (C18) |  |  |  |  |  |  |  |  |  |
| Cases | 118 | 87 | 102 | 114 |  | 421 | | | |
| Model 0, HR (95% CI) | 1.00 (0.83-1.20) | 0.71 (0.58-0.88) | 0.82 (0.68-1.00) | 0.92 (0.77-1.11) |  | 1.00 (0.89-1.12) | 0.9623 | 1.00 (0.88-1.12) | 0.9602 |
| Model 1, HR (95% CI) | 1.00 (0.82-1.22) | 0.77 (0.62-0.94) | 0.94 (0.77-1.14) | 1.10 (0.90-1.35) |  | 1.08 (0.95-1.22) | 0.2353 | 1.08 (0.95-1.23) | 0.2368 |
| Model 2, HR (95%CI) | 1.00 (0.81-1.23) | 0.79 (0.64-0.97) | 0.99 (0.81-1.20) | 1.20 (0.97-1.48) |  | 1.12 (0.98-1.28) | 0.0911 | 1.13 (0.98-1.29) | 0.0920 |
| Rectum (C19-20) |  |  |  |  |  |  |  |  |  |
| Cases | 33 | 37 | 39 | 45 |  | 154 | | | |
| Model 0, HR (95% CI) | 1.00 (0.71-1.41) | 1.10 (0.80-1.52) | 1.16 (0.84-1.58) | 1.32 (0.99-1.77) |  | 1.12 (0.94-1.35) | 0.2108 | 1.13 (0.93-1.38) | 0.2105 |
| Model 1, HR (95% CI) | 1.00 (0.69-1.45) | 1.03 (0.75-1.42) | 1.04 (0.76-1.43) | 1.17 (0.85-1.60) |  | 1.07 (0.87-1.31) | 0.5134 | 1.08 (0.87-1.34) | 0.5124 |
| Model 2, HR (95%CI) | 1.00 (0.68-1.47) | 1.09 (0.79-1.50) | 1.16 (0.85-1.58) | 1.39 (1.00-1.94) |  | 1.15 (0.93-1.43) | 0.1904 | 1.16 (0.93-1.46) | 0.1899 |
| Liver (C22) |  |  |  |  |  |  |  |  |  |
| Cases | 11 | 13 | 7 | 18 |  | 49 | | | |
| Model 0, HR (95% CI) | 1.00 (0.55-1.81) | 1.14 (0.66-1.97) | 0.62 (0.30-1.30) | 1.59 (1.00-2.53) |  | 1.21 (0.87-1.67) | 0.2575 | 1.22 (0.87-1.72) | 0.2518 |
| Model 1, HR (95% CI) | 1.00 (0.53-1.90) | 1.25 (0.73-2.15) | 0.74 (0.35-1.54) | 1.82 (1.07-3.10) |  | 1.28 (0.89-1.83) | 0.1820 | 1.30 (0.89-1.90) | 0.1777 |
| Model 2, HR (95%CI) | 1.00 (0.52-1.93) | 1.24 (0.72-2.12) | 0.71 (0.34-1.48) | 1.61 (0.91-2.84) |  | 1.21 (0.83-1.76) | 0.3240 | 1.23 (0.82-1.83) | 0.3174 |
| Pancreas (C25) |  |  |  |  |  |  |  |  |  |
| Cases | 29 | 34 | 30 | 30 |  | 123 | | | |
| Model 0, HR (95% CI) | 1.00 (0.69-1.44) | 1.14 (0.81-1.59) | 1.01 (0.70-1.44) | 1.00 (0.70-1.43) |  | 0.98 (0.79-1.21) | 0.8486 | 0.98 (0.78-1.22) | 0.8507 |
| Model 1, HR (95% CI) | 1.00 (0.67-1.49) | 1.19 (0.85-1.66) | 1.09 (0.76-1.55) | 1.10 (0.75-1.62) |  | 1.02 (0.81-1.29) | 0.8688 | 1.02 (0.80-1.30) | 0.8668 |
| Model 2, HR (95%CI) | 1.00 (0.67-1.50) | 1.19 (0.86-1.67) | 1.08 (0.75-1.54) | 1.10 (0.73-1.64) |  | 1.02 (0.80-1.30) | 0.8875 | 1.02 (0.79-1.32) | 0.8852 |
| Lung (C34) |  |  |  |  |  |  |  |  |  |
| Cases | 105 | 89 | 107 | 106 |  | 407 | | | |
| Model 0, HR (95% CI) | 1.00 (0.83-1.21) | 0.85 (0.69-1.05) | 1.02 (0.85-1.24) | 1.03 (0.85-1.25) |  | 1.04 (0.93-1.16) | 0.5279 | 1.04 (0.92-1.17) | 0.5324 |
| Model 1, HR (95% CI) | 1.00 (0.81-1.24) | 0.81 (0.66-1.00) | 0.96 (0.79-1.15) | 0.90 (0.73-1.10) |  | 0.98 (0.86-1.11) | 0.7397 | 0.98 (0.85-1.12) | 0.7333 |
| Model 2, HR (95%CI) | 1.00 (0.80-1.24) | 0.85 (0.69-1.04) | 1.04 (0.86-1.25) | 1.02 (0.83-1.27) |  | 1.04 (0.91-1.18) | 0.5933 | 1.04 (0.90-1.20) | 0.5997 |
| Melanoma (C43) |  |  |  |  |  |  |  |  |  |
| Cases | 66 | 62 | 57 | 81 |  | 266 | | | |
| Model 0, HR (95% CI) | 1.00 (0.79-1.27) | 0.91 (0.71-1.17) | 0.84 (0.65-1.09) | 1.20 (0.96-1.49) |  | 1.09 (0.95-1.26) | 0.2255 | 1.10 (0.95-1.27) | 0.2221 |
| Model 1, HR (95% CI) | 1.00 (0.77-1.30) | 0.86 (0.67-1.10) | 0.78 (0.60-1.01) | 1.10 (0.87-1.40) |  | 1.06 (0.91-1.24) | 0.4421 | 1.07 (0.91-1.26) | 0.4360 |
| Model 2, HR (95%CI) | 1.00 (0.76-1.32) | 0.86 (0.67-1.10) | 0.77 (0.59-0.99) | 1.05 (0.82-1.35) |  | 1.04 (0.88-1.23) | 0.6211 | 1.05 (0.88-1.24) | 0.6136 |
| Meothelioma (C45) |  |  |  |  |  |  |  |  |  |
| Cases | 4 | 5 | 8 | 6 |  | 23 | | | |
| Model 0, HR (95% CI) | 1.00 (0.37-2.67) | 1.13 (0.47-2.70) | 1.81 (0.90-3.62) | 1.36 (0.61-3.02) |  | 1.15 (0.71-1.86) | 0.5598 | 1.16 (0.70-1.93) | 0.5644 |
| Model 1, HR (95% CI) | 1.00 (0.35-2.83) | 0.85 (0.34-2.11) | 1.32 (0.66-2.64) | 0.83 (0.36-1.94) |  | 0.95 (0.56-1.62) | 0.8481 | 0.94 (0.54-1.66) | 0.8418 |
| Model 2, HR (95%CI) | 1.00 (0.34-2.97) | 0.86 (0.35-2.14) | 1.35 (0.68-2.69) | 0.83 (0.33-2.05) |  | 0.95 (0.54-1.68) | 0.8571 | 0.94 (0.52-1.72) | 0.8502 |
| Breast (C50) |  |  |  |  |  |  |  |  |  |
| Cases | 539 | 524 | 425 | 382 |  | 1870 | | | |
| Model 0, HR (95% CI) | 1.00 (0.92-1.09) | 0.96 (0.88-1.04) | 0.78 (0.71-0.86) | 0.71 (0.64-0.78) |  | **0.85 (0.80-0.90)** | **<0.0001** | **0.84 (0.79-0.89)** | **<0.0001** |
| Model 1, HR (95% CI) | 1.00 (0.91-1.10) | 0.99 (0.91-1.08) | 0.84 (0.76-0.92) | 0.78 (0.70-0.87) |  | **0.89 (0.84-0.94)** | **0.0002** | **0.88 (0.83-0.94)** | **0.0002** |
| Model 2, HR (95%CI) | 1.00 (0.91-1.10) | 1.00 (0.92-1.09) | 0.86 (0.78-0.94) | 0.83 (0.74-0.92) |  | **0.91 (0.85-0.97)** | **0.0039** | **0.91 (0.85-0.97)** | **0.0041** |
| Endometrial (C54.1) |  |  |  |  |  |  |  |  |  |
| Cases | 147 | 104 | 66 | 55 |  | 372 | | | |
| Model 0, HR (95% CI) | 1.00 (0.85-1.18) | 0.69 (0.57-0.83) | 0.44 (0.34-0.55) | 0.36 (0.28-0.47) |  | **0.62 (0.54-0.71)** | **<0.0001** | **0.60 (0.52-0.69)** | **<0.0001** |
| Model 1, HR (95% CI) | 1.00 (0.83-1.20) | 0.85 (0.71-1.03) | 0.64 (0.50-0.82) | 0.61 (0.46-0.81) |  | **0.79 (0.68-0.91)** | **0.0013** | **0.78 (0.67-0.91)** | **0.0014** |
| Model 2, HR (95%CI) | 1.00 (0.83-1.21) | 0.86 (0.72-1.04) | 0.65 (0.50-0.83) | 0.61 (0.46-0.82) |  | **0.79 (0.68-0.92)** | **0.0023** | **0.78 (0.66-0.92)** | **0.0024** |
| Ovarian (C56) |  |  |  |  |  |  |  |  |  |
| Cases | 49 | 60 | 54 | 66 |  | 229 | | | |
| Model 0, HR (95% CI) | 1.00 (0.76-1.32) | 1.20 (0.93-1.54) | 1.08 (0.82-1.40) | 1.32 (1.04-1.69) |  | 1.11 (0.95-1.29) | 0.1936 | 1.11 (0.95-1.31) | 0.1912 |
| Model 1, HR (95% CI) | 1.00 (0.74-1.35) | 1.17 (0.91-1.51) | 1.06 (0.82-1.39) | 1.32 (1.02-1.71) |  | 1.11 (0.94-1.31) | 0.2274 | 1.12 (0.94-1.33) | 0.2245 |
| Model 2, HR (95%CI) | 1.00 (0.73-1.36) | 1.15 (0.89-1.48) | 1.01 (0.78-1.32) | 1.22 (0.93-1.60) |  | 1.07 (0.90-1.28) | 0.4489 | 1.08 (0.89-1.29) | 0.4441 |
| Kidney (C64-65) |  |  |  |  |  |  |  |  |  |
| Cases | 35 | 27 | 20 | 20 |  | 102 | | | |
| Model 0, HR (95% CI) | 1.00 (0.72-1.39) | 0.75 (0.52-1.10) | 0.55 (0.36-0.86) | 0.57 (0.36-0.88) |  | **0.76 (0.60-0.97)** | **0.0285** | **0.75 (0.58-0.97)** | **0.0289** |
| Model 1, HR (95% CI) | 1.00 (0.69-1.44) | 0.75 (0.52-1.09) | 0.58 (0.37-0.90) | 0.60 (0.37-0.95) |  | 0.79 (0.60-1.02) | 0.0754 | 0.78 (0.59-1.03) | 0.0763 |
| Model 2, HR (95%CI) | 1.00 (0.68-1.46) | 0.75 (0.52-1.09) | 0.59 (0.38-0.91) | 0.62 (0.38-1.01) |  | 0.80 (0.61-1.06) | 0.1184 | 0.79 (0.59-1.06) | 0.1196 |
| Bladder (C67) |  |  |  |  |  |  |  |  |  |
| Cases | 12 | 12 | 15 | 18 |  | 57 | | | |
| Model 0, HR (95% CI) | 1.00 (0.57-1.76) | 0.94 (0.53-1.65) | 1.19 (0.71-1.97) | 1.45 (0.91-2.30) |  | 1.21 (0.89-1.63) | 0.2278 | 1.22 (0.88-1.68) | 0.2282 |
| Model 1, HR (95% CI) | 1.00 (0.54-1.86) | 1.01 (0.57-1.78) | 1.28 (0.78-2.12) | 1.52 (0.91-2.53) |  | 1.22 (0.87-1.71) | 0.2449 | 1.23 (0.87-1.76) | 0.2457 |
| Model 2, HR (95%CI) | 1.00 (0.53-1.90) | 1.05 (0.60-1.87) | 1.41 (0.86-2.31) | 1.66 (0.97-2.83) |  | 1.26 (0.89-1.80) | 0.1967 | 1.28 (0.88-1.86) | 0.1977 |
| Brain (C71) |  |  |  |  |  |  |  |  |  |
| Cases | 19 | 18 | 20 | 17 |  | 74 | | | |
| Model 0, HR (95% CI) | 1.00 (0.64-1.57) | 0.91 (0.58-1.45) | 1.02 (0.66-1.58) | 0.88 (0.55-1.41) |  | 0.96 (0.73-1.26) | 0.7690 | 0.96 (0.72-1.28) | 0.7665 |
| Model 1, HR (95% CI) | 1.00 (0.61-1.63) | 0.89 (0.56-1.41) | 0.99 (0.64-1.53) | 0.89 (0.54-1.48) |  | 0.97 (0.72-1.31) | 0.8300 | 0.97 (0.70-1.33) | 0.8275 |
| Model 2, HR (95%CI) | 1.00 (0.60-1.67) | 0.88 (0.56-1.40) | 0.95 (0.62-1.48) | 0.82 (0.48-1.40) |  | 0.93 (0.68-1.28) | 0.6697 | 0.93 (0.66-1.30) | 0.6672 |
| Non-Hodgkin Lymphoma (C82-85) | |  |  |  |  |  |  |  |  |
| Cases | 46 | 47 | 58 | 59 |  | 210 | | | |
| Model 0, HR (95% CI) | 1.00 (0.75-1.34) | 0.97 (0.73-1.28) | 1.20 (0.93-1.55) | 1.21 (0.94-1.57) |  | 1.11 (0.94-1.30) | 0.2092 | 1.11 (0.94-1.32) | 0.2113 |
| Model 1, HR (95% CI) | 1.00 (0.73-1.37) | 0.94 (0.71-1.25) | 1.15 (0.89-1.49) | 1.15 (0.87-1.51) |  | 1.08 (0.91-1.29) | 0.3802 | 1.09 (0.90-1.31) | 0.3836 |
| Model 2, HR (95%CI) | 1.00 (0.72-1.39) | 0.96 (0.72-1.27) | 1.17 (0.91-1.51) | 1.14 (0.86-1.52) |  | 1.08 (0.90-1.29) | 0.4309 | 1.08 (0.89-1.31) | 0.4351 |
| Diffuse non-Hodgkins lymphoma (C83) | |  |  |  |  |  |  |  |  |
| Cases | 30 | 18 | 30 | 28 |  | 106 | | | |
| Model 0, HR (95% CI) | 1.00 (0.70-1.43) | 0.57 (0.36-0.90) | 0.94 (0.66-1.35) | 0.87 (0.60-1.27) |  | 1.01 (0.81-1.26) | 0.9393 | 1.01 (0.79-1.28) | 0.9473 |
| Model 1, HR (95% CI) | 1.00 (0.67-1.49) | 0.55 (0.35-0.88) | 0.91 (0.64-1.30) | 0.85 (0.57-1.26) |  | 1.00 (0.78-1.28) | 0.9985 | 1.00 (0.77-1.30) | 0.9930 |
| Model 2, HR (95%CI) | 1.00 (0.66-1.52) | 0.59 (0.37-0.93) | 0.99 (0.70-1.41) | 0.95 (0.62-1.43) |  | 1.05 (0.81-1.36) | 0.7393 | 1.05 (0.79-1.38) | 0.7482 |
| Multiple Myeloma (C90) | |  |  |  |  |  |  |  |  |
| Cases | 24 | 17 | 20 | 21 |  | 82 | | | |
| Model 0, HR (95% CI) | 1.00 (0.67-1.49) | 0.69 (0.43-1.11) | 0.81 (0.52-1.25) | 0.86 (0.56-1.32) |  | 0.97 (0.75-1.25) | 0.7898 | 0.96 (0.73-1.26) | 0.7883 |
| Model 1, HR (95% CI) | 1.00 (0.64-1.55) | 0.79 (0.49-1.26) | 1.05 (0.68-1.63) | 1.28 (0.80-2.03) |  | 1.15 (0.87-1.52) | 0.3374 | 1.16 (0.86-1.56) | 0.3390 |
| Model 2, HR (95%CI) | 1.00 (0.63-1.58) | 0.78 (0.49-1.26) | 1.04 (0.67-1.61) | 1.26 (0.77-2.06) |  | 1.14 (0.85-1.54) | 0.3809 | 1.15 (0.84-1.58) | 0.3826 |
| Leukemia (C91-95) |  |  |  |  |  |  |  |  |  |
| Cases | 33 | 22 | 28 | 31 |  | 114 | | | |
| Model 0, HR (95% CI) | 1.00 (0.71-1.41) | 0.63 (0.42-0.96) | 0.80 (0.55-1.16) | 0.88 (0.62-1.25) |  | 0.99 (0.79-1.23) | 0.9083 | 0.99 (0.78-1.24) | 0.9058 |
| Model 1, HR (95% CI) | 1.00 (0.69-1.46) | 0.63 (0.42-0.96) | 0.80 (0.56-1.16) | 0.89 (0.61-1.30) |  | 0.99 (0.78-1.26) | 0.9606 | 0.99 (0.77-1.28) | 0.9578 |
| Model 2, HR (95%CI) | 1.00 (0.68-1.48) | 0.63 (0.42-0.96) | 0.80 (0.55-1.15) | 0.87 (0.59-1.30) |  | 0.99 (0.77-1.27) | 0.9158 | 0.99 (0.76-1.28) | 0.9130 |

Estimates bold if associations are statistically significant (p<0.05).

Model 0: Stratified by age group, geographical region and adjusted for Townsend deprivation score

Model 1: Model 0 + adjusted for racial/ethnic group, height, lives with a spouse or partner, body mass index, cigarette smoking, alcohol consumption, total physical activity, hormone replacement therapy use, oral contraceptive use, and parity and age at first birth

Model 2: Model 1 + additionally adjusted for serum concentrations of insulin-like growth factor-I, C-reactive protein, glycated hemoglobin (fourths, unknown).

Abbreviations: CI=Confidence interval; HR=hazard ratio; IQR=interquartile range; SHBG=sex hormone binding globulin.

Supplementary Table S7: HR and 95% CIs for malignant melanoma in men after correction for factors relating to sun exposure

| Hormone | Quartiles | N cases | HR (95% CI) | P_trend_ |
| --- | --- | --- | --- | --- |
| Free testosterone | 1 | 109 | 1.00(0.82-1.21) | |
|  | 2 | 109 | 1.12(0.92-1.35) | |
|  | 3 | 143 | 1.65(1.40-1.94) | |
|  | 4 | 108 | 1.53(1.26-1.86) | |
|  | per 50 pmol/L |  | 1.36(1.15-1.62) | 0.0004 |
| Total testosterone | 1 | 124 | 1.00(0.83-1.20) | |
|  | 2 | 127 | 1.07(0.90-1.28) | |
|  | 3 | 131 | 1.17(0.98-1.38) | |
|  | 4 | 140 | 1.36(1.15-1.61) | |
|  | per 5 nmol/L |  | 1.29(1.06-1.57) | 0.01 |
| SHBG | 1 | 98 | 1.00(0.81-1.23) | |
|  | 2 | 124 | 1.09(0.92-1.30) | |
|  | 3 | 128 | 1.05(0.89-1.25) | |
|  | 4 | 122 | 0.98(0.81-1.18) | |
|  | per 10 nmol/L | 109 | 0.98(0.91-1.06) | 0.70 |

†Associations stratified for age group, geographical region and adjusted for Townsend deprivation score, racial/ethnic group, height, lives with a spouse or partner, body mass index, cigarette smoking, alcohol consumption, total physical activity, skin color, hair color, skin reaction to sun exposure, and sunburn before age 15, and corrected for regression dilution bias.

Abbreviations: CI=Confidence interval; HR=hazard ratio; SHBG= sex hormone binding globulin.
